# A Sequential Multiple Assignment Randomized Trial Design with Response-Adaptive Tailoring Function

**DOI:** 10.64898/2026.04.28.26351992

**Authors:** Zhengxi Chen, Holly Hartman

## Abstract

We present a novel sequential multiple assignment randomized trial (SMART) design that integrates response-adaptive randomization with tailoring functions (RA-TF-SMART). We develop percentile-based and Z-score RA-TFs that incorporate both within-patient and between-patient adaptation to map continuous outcomes to randomization probabilities. We apply Q-learning, tree-based reinforcement learning, and G-estimation to estimate dynamic treatment regimens (DTRs). We compare our RA-TF-SMART designs to balanced randomized SMARTs (BR-SMARTs), tailoring function SMARTs (TF-SMARTs), and generalized outcome-adaptive SMARTs (GO-SMARTs). This study addresses limitations in SMART methodology by presenting designs where randomization probability does not require dichotomization of continuous outcomes and utilizes both individual patient outcomes and accumulated treatment efficacy data from prior participants. RA-TF-SMARTs offer a flexible framework that maximizes benefit for trial participants while maintaining statistical validity for post-trial DTR estimation.

## 1. Introduction

Dynamic treatment regimens (DTRs) are prespecified, sequential decision-making rules designed to ensure that patients are provided with effective treatment at the appropriate time. The necessity of DTRs arises from the heterogeneity in treatment responses across patients, the changing effectiveness of treatments over time due to evolving risk factors and fluctuations in health status, potential for relapse, as well as considerations of treatment costs, side effects, and adherence challenges.^1-5^ DTRs consist of four main components: the decision point, the tailoring variable, the intervention options, and the decision rule.^2,6^ Firstly, decision points are time points when treatment adaptations occur. The tailoring variable captures patient information that is used to identify the optimal treatment decisions at subsequent stages. Interventions can be adjusted in terms of the types of treatments, treatment combinations, intensity, dosage, delivery methods, strategies to improve engagement and adherence, or by continuing with the same treatment as in earlier stages. Lastly, decision rules are predefined criteria used to connect tailoring variables to intervention options at each decision point. By adapting treatment strategies to individual patient characteristics and intermediate responses, DTRs can significantly enhance resource utilization and intervention effectiveness.^6^ The conceptual advantages of sequential treatments and DTRs over fixed-intervention methods were first recognized by behavioral and psychiatric scientists. DTRs have been applied to study major depressive disorder,^7^ child and adolescent mental health,^8^ Alcohol use disorder,^9^ Attention Deficit/Hyperactivity Disorder (ADHD)^10^ and Autism Spectrum Disorder.^11^ Recently, an increasing number of studies have implemented DTRs to other medical fields, including chronic diseases and oncology.^12,13,14,15^

Sequential, multiple assignment, randomized trials (SMARTs) provide a systematic framework for constructing and evaluating DTRs.^1,2,16-18^ SMARTs are multi-stage, sequential randomized trials in which participants are followed through different treatment stages and assigned to predetermined treatment options at the beginning of each stage either randomly or deterministically depending on response to previous treatments. Each stage corresponds to one of the decision points in a DTR. Similar to conventional single-stage trials, the randomization in SMARTs enables researchers to make statistically valid causal inferences about the efficacy of different intervention options without relying on unprovable assumptions as is the case in many observational studies.^19,20^ SMARTs have been rapidly accepted within the clinical and health services research community, with applications spanning chronic diseases such as obesity and HIV, and psychiatric and mental health disorders such as schizophrenia, depression, and ADHD.^2,3,21-27^ The aim of a SMART is to identify the DTR that optimizes the expected outcome at the end of the final stage of treatment, which requires determining the optimal set of decision points, decision rules, treatment options, and tailoring variables.^21,26^

Existing SMART designs face important limitations that may restrict their widespread adoption. The Balanced Randomization SMART (BR-SMART) is the most simplified version of the SMART design, which assigns participants to available treatments equally at each stage. While BR-SMART ensures equal distribution across all DTRs and maximizes statistical power when comparing different DTRs, it mainly serves as a methodological benchmark for comparing and measuring the effectiveness of other SMART designs due to not incorporating participants’ response.^16^ Tailoring variable SMARTs (TV-SMARTs) are the most widely implemented SMART design for evaluating DTRs, where treatment decisions after the first stage are guided by predefined tailoring variables. These tailoring variables generally follow a binary rule to reflect response versus non-response to previous treatments. For example, in a previous SMART study, all overweight or obese adults received short-duration Intensive Behavioral Therapy (IBT) in the first stage.^26^ After five weeks, patients who successfully lost five pounds or more were considered as responders and continued with the standard IBT, while non-responders were transitioned to an augmented IBT arm that incorporated meal replacements. In this case, five-pound weight loss at week five serves as the binary tailoring variable to guide the treatment randomization in the second stage. A notable limitation of the TV-SMARTs framework is that it only accounts for within-patient treatment adjustments, without considering adaptations between patients. This oversight emphasizes the necessity for more advanced strategies considering between-patient variation in response to treatments.

To better utilize information from previous patients in the same trial, response-adaptive randomization (RAR) has been proposed to update randomization probabilities continuously, favoring more effective treatments based on accumulating treatment responses as more patients are enrolled in the trial.^28-30^ This dynamic allocation approach offers significant ethical and cost advantages by limiting patient exposure to ineffective treatments and increasing efficiency through early termination of futile treatments.^31,32^ Recently, Wang et al. introduced a response-adaptive SMART (RA-SMART) design that shifts the second-stage randomization probabilities toward more effective treatments based on accumulated stage-one response data from earlier patients.^33^ Yang et al. proposed a generalized outcome-adaptive SMART (GO-SMART) design, which adapts second-stage randomization probability based on patient’s response to first-stage treatment and conditional probability of responding to intervention options in stage two.^34^ One critical limitation shared by both RA-SMART and GO-SMART is their exclusive focus on binary response indicators and binary outcomes, which restricts their applicability in studies with continuous intermediate and final outcomes. In many studies, the outcome of interest may not be naturally binary, requiring dichotomization of continuous outcomes to create a binary response indicator. This dichotomization introduces several limitations, including loss of information and statistical power, increased risk of misclassification and bias, underestimation of variability, and masking of non-linear relationships.^35,36^ Additionally, the selection of cutoff points is often inconsistent across studies, as it is typically arbitrary and lacks a standardized approach, making it challenging to identify objectively optimal cutoff points for most intermediate outcomes.^37^

To estimate DTRs more flexibly without defining binary response indicator, Hartman et al. proposed a SMART design in which the second-stage treatment is determined based on a tailoring function (TF).^38,39^ TFs extend the concept of tailoring variables by using a continuous outcome at the end of the first stage to calculate the randomization probabilities in the next stage following a multinomial distribution. This approach preserves the information contained in continuous outcomes and avoids arbitrary dichotomization. However, the TFs proposed rely on direct linear transformations that require pre-specification of outcome ranges and assume outcomes are appropriately scaled relative to a fixed denominator.^38^ TF-SMARTs may perform sub-optimally when outcome distributions are not centered around the transformation midpoint and lead to systematic over- or under-allocation to treatment switching regardless of relative performance. Additionally, the TF-SMART framework only accounts for within-patient treatment adjustments, without considering between-patient adaptation that could benefit trial participants by utilizing accumulating efficacy data from previous patients. That is, if a person is randomized to switch treatments, TF-SMARTs assign balanced randomization probabilities to available treatments and cannot favor one based on accumulating patient data.

In this study, we propose a Response-Adaptive Tailoring Function SMART (RA-TF-SMART) design that incorporates both within-patient and between-patient adaptation with continuous outcomes measured at each decision point. We aim to allocate more participants to superior DTRs based on accumulating patient data while maintaining efficient DTR evaluation. In Section 2, we introduce the DTR framework, notation and SMART designs. In Section 3, we present our weighting strategy and methods for evaluating DTRs. Our simulation studies are introduced in Section 4. We report simulation results in Section 5 and end with a discussion in Section 6.

## 2. Method

### 2.1 DTR Notation, SMART Design and Assumptions

Interventions for DTRs can be depicted as a three-part structure that outlines the sequence of recommended interventions in the order of first-stage treatment, second-stage treatment for responders, and second-stage treatment for non-responders. In this study, we concentrate on DTRs embedded in two-stage SMARTs as this is the most common design currently in use. However, SMARTs can be extended to include more than two decision points. For simplicity, the designs in this study only offer two intervention options at each randomization point, but the methods provided here can be extended to three or more intervention options at any stage. **Figure 1** presents the randomization structures of the four SMART designs evaluated in this study: (I) BR-SMART, (II) TF-SMART, (III) GO-SMART, and (IV) the proposed RA-TF-SMART. All designs embed four DTRs: AAC, AAD, BBC, and BBD.

**Figure 1.**
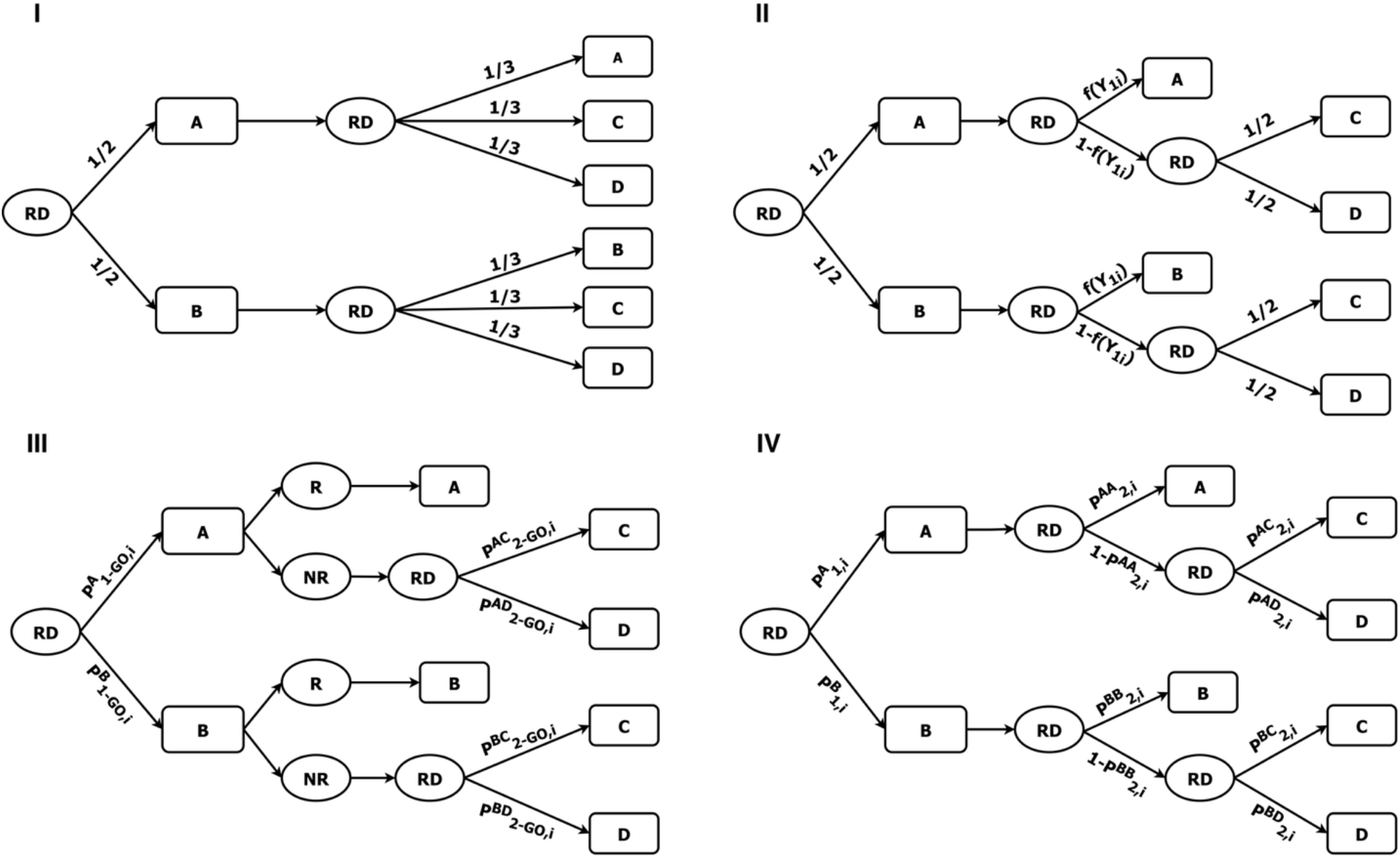
Schematic of two-stage sequential multiple assignment randomized trial (SMART) designs: (I) balanced randomization SMART, (II) tailoring function SMART, (III) generalized outcome adaptive SMART, and (IV) response-adaptive tailoring function SMART. A, B, C and D represent treatments, RD indicates a randomization point, R and NR represent response and non-response, and the expressions along the lines indicate the randomization probability of the treatment defined in Section 2.

Let *T*_1_ denote the first-stage treatment with available treatment options *j* ∈ 𝒥 = {1, …, *J*} and *T*_2_ denote the second-stage treatment with available treatment options *l* ∈ ℒ_*j*_ = {1, …, *L*_*j*_} conditional on first-stage treatment *j*. We assume a linear relationship between the second-stage outcome *Y*_2_ and the first-stage outcome *Y*_1_ conditional on treatment sequence *d*(*T*_1_, *T*_2_). Specifically, for patients assigned to first-stage treatment *j* and second-stage treatment *l*, the expected second-stage outcome is modeled as *E*(*Y*_2_|*Y*_1_, *T*_1_ = *j, T*_2_ = *l*) = *β*_*jl*_ + *γ*_*jl*_*Y*_1_, where *β*_*jl*_ represents the intercept and *γ*_*jl*_ represents the slope for treatment sequence *d*(*j, l*). In addition, we assume that higher *Y*_1_ values correspond to superior treatment performance; however, this design can be adapted to accommodate outcomes where lower values indicate better performance.

### 2.2 Tailoring Function

TFs are a generalization of tailoring variables designed to map a patient’s intermediate outcome to produce valid second-stage randomization probabilities.^38,39^ Specifically, the tailoring function, denoted as *f*(*Y*_1_), maps the first-stage outcome, *Y*_1_, to the interval [0, 1], where the resulting value represents the probability of continuing with the same treatment. Patients with more favorable outcomes receive higher probabilities of continuing their current treatment. The selection of an appropriate TF depends on the characteristics of the outcome variable, and various cumulative distribution functions may be considered. A critical characteristic of TF is that it ensures representation in all treatment sequences by the end of the trial, allowing for the estimation of all treatment pathway effects and maintaining the positivity assumption. Detailed descriptions of examples and power transformations of TFs, as well as the construction of TF-SMARTs are provided in Hartman et al.^38^

### 2.3 Generalized Outcome-Adaptive SMART

To apply the GO-SMART scheme for continuous outcomes, continuous outcomes *Y* ∈ {*Y*_1_, *Y*_2_} are dichotomized into binary response indicators *R* ∈ {*R*_1_, *R*_2_} using pre-specified thresholds defined separately for each stage, with values 1 for response and 0 for non-response. This dichotomization is expressed as *R*_1*i*_ = *I*(*Y*_1*i*_ > *φ*_1_) and *R*_2*i*_ = *I*(*Y*_2*i*_ > *φ*_2_) for patient *i*, where *φ*_1_ and *φ*_2_ denote the response thresholds for stages one and two, respectively. In this study, we implement GO-SMART AR-1 as recommended by Yang et al.^34^

#### 2.3.1 Response Definition

The stage one response rate is defined as 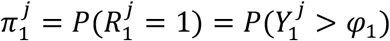, where 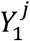 represents the continuous outcome measured at the end of stage one for patients treated with *j* ∈ 𝒥, and 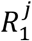 represents the corresponding binary response indicator. The conditional response rate for stage two is defined as 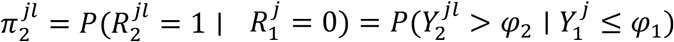, where 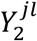 denotes the continuous outcome observed at the end of stage two for a patient who received treatment *l* (where *l* ≠ *j*) in the second stage, following an observed non-response to treatment *j* in stage one, and 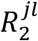 represents the second-stage binary response indicator.

#### 2.3.2 Burn-in Period

Let *p*_0_ denote a pre-specified burn-in proportion and define *n*_0_ = ⌈*p*_0_*n*⌉ < *n* as the burn-in sample where *n* is the total sample size. During the burn-in period, we employ balanced randomization to ensure equal representation across all treatment sequences. In the first stage, each patient *i* = 1, …, *n*_0_ is randomized to treatment option *j* with equal probability 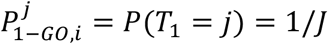 for all *j* ∈ 𝒥 = {1, …, *J*}. Similarly in the second stage, patients are randomized to treatment option *l* with equal probability 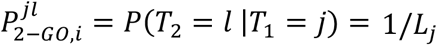 for all *l* ∈ ℒ_*j*_ = {1, …, *L*_*j*_} given their first-stage treatment *j*. Under *φ*_1_ and *φ*_2_, their corresponding first-stage response, *R*_1*i*_, and second-stage response, *R*_2*i*_, are determined for each patient *i* at the end of each stage.

#### 2.3.3 Stage-one Adaptation Using Binary Response

Subsequent patients (*i* = *n*_0_ + 1, …, *n*) are randomized to treatment *j* ∈ 𝒥 with first-stage randomization probability defined as 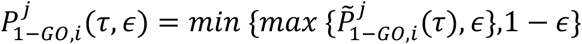 and 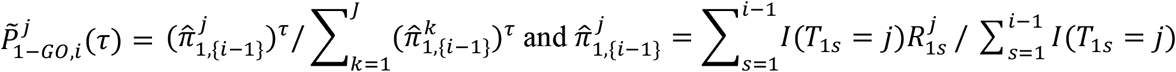 represents the empirical stage one response rate for treatment *j* based on data from the first *i* − 1 patients. The tuning parameter *τ* ∈ [0, ∞) controls the degree of adaptation based on the accumulated response rates. When *τ* = 1, randomization probabilities are proportional to the observed response rates, *τ* > 1 favor randomization toward treatments with higher response rates, while *τ* < 1 moderates this imbalance toward equal randomization. The constraint parameter *ϵ* ∈ (0,1) bounds extreme randomization probabilities away from 0 and 1 to ensure positivity for valid causal inference.

#### 2.3.4 Stage-two Adaptation Using Binary Response

Patients who respond (i.e., 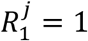) to *T*_1_ = *j* are assigned to *j* in the second stage. For patients who experience non-response (i.e., 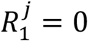) in stage one, patients are assigned to treatment *l* (*l* ≠ *j*) in stage two according to adaptive probability 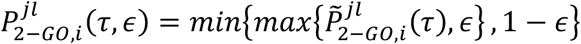. GO-SMART AR-1 is a conditional response-based approach with second-stage randomization probability defined as 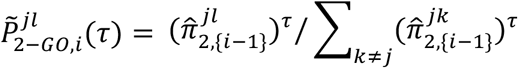, where 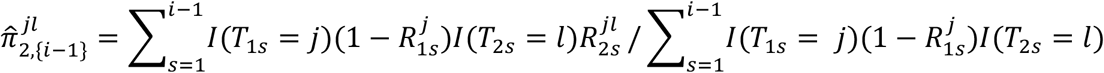 denotes the empirical conditional stage two response rate for *l* given non-response to *j*.

### 2.4 Response-Adaptive Tailoring Function SMART

#### 2.4.1 Stage-one Adaptation Using Continuous Outcome

Following an initial burn-in period identical to the method used in the GO-SMART design (**Section 2.3.2**), patient *i* = *n*_0_ + 1, …, *n* is randomized to treatment *j* in stage one with probability 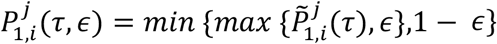, where 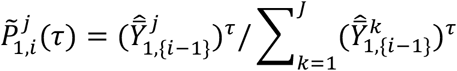 and 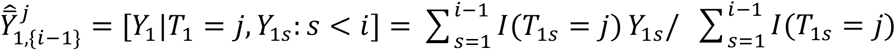 represents the empirical expectation of *Y*_1_ for treatment *j* based on the first *i* − 1 patients. As in the GO-SMART design, the tuning parameter *τ* controls the degree of adaptation intensity while *ϵ* bounds probabilities within [*ϵ*, 1 − *ϵ*] to avoid extreme randomization probability.

#### 2.4.2 Stage-two Adaptation Using Continuous Outcome

For patient *i* = *n*_0_ + 1, …, *n* assigned to treatment *j* in stage 1, the second-stage randomization involves two sequential decisions: (1) between continuation of *T*_1_ = *j* and switching to another treatment, and (2) between two alternative treatments among those who switch treatments. The second-stage randomization is determined by the observed *Y*_1_s of the *i*^*th*^ patient and the previous *i* − 1 patients who received the same *T*_1_ as the *i*^*th*^ patient through RA-TFs.

##### 2.4.2.1 Adaptive Cutoff Estimation

To maintain appropriate randomization probabilities relative to the decision rules between treatment sequences, we incorporate an adaptive cutoff estimation mechanism to implement distributional centering for RAR. Consider a scenario where *Y*_1_ has a population mean of 40 and the true optimal cutoff falls at 20. Without distributional centering, patients with *Y*_1_ between 20 and 40 would receive probabilities below 0.5 for treatment continuation despite falling above the optimal threshold. This adaptive cutoff-updating mechanism automatically adjusts randomization probabilities to reflect the emerging relationship between outcome distributions and treatment benefit, which is particularly valuable when outcome distribution differs from prior knowledge or when implementing RAR across diverse populations with varying *Y*_1_.

The optimal cutoff 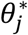 for each *T*_1_, represents the *Y*_1_ value where the expected benefit of continuing treatment *j* equals the expected benefit of switching. This cutoff is estimated using weighted regression on the previous *i* − 1 patients who received *j* in stage one. Observations are weighted by inverse probability of treatment weighting (IPTW) to account for adaptive randomization.^40,41^ The weight, *w* = 1/*P*(*T*_2_ = *l* ∣ *T*_1_ = *j*), is constructed as the inverse of the probability of receiving observed treatment sequence *d*(*j, l*). Using only the data of patients with *T*_1_ = *j*, we fit the weighted linear model that simultaneously captures the cutoffs between all *T*_2_ options:

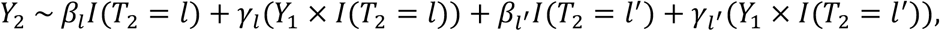

where *I*(*T*_2_ = *l*) and *I*(*T*_2_ = *l*′) are indicators for switching to treatments *l* and *l*′ ∈ ℒ_*j*_ (*l* ≠ *l*′ ≠ *j*), respectively. The reference category (when both *I*(*T*_2_ = *l*) = 0 and *I*(*T*_2_ = *l*′) = 0) represents continuation with treatment *j*. The cutoff between continuing *j* and switching to *l* is defined as 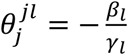. Similarly, the cutoff between continuing *j* and switching to *l*′ is 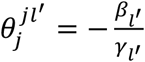. The gate cutoff is defined as 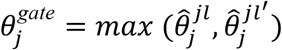 which determines the decision of continuing or switching. For patients who are randomized to switch treatments, the allocation between *l* and *l*′ is guided by an additional cutoff 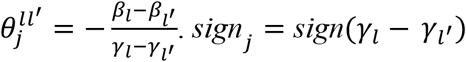 indicates superiority between *l* and *l*′ relative to the cutoff 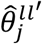. That is, when 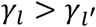 (i.e. when sign_*j*_ = +1), treatment *l* is superior to *l*′ when 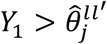.

Cutoff estimates can fluctuate substantially between consecutive patients due to sampling variability, especially during early enrollment when sample sizes within treatment arms are small. To stabilize the cutoff estimation and ensure smooth randomization adaptation, we implement the exponential moving average (EMA) update rule^42^ defined as 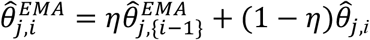, where 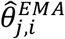 denotes the adjusted cutoff estimate after EMA update, 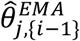 represents the estimate after EMA update based on previous *i* − 1 patients in treatment arm 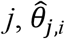 is the estimate incorporating the current patient *i*, and *η* ∈ [0,1] is the smoothing parameter controlling the weight assigned to historical information. For simplicity, we denote the adjusted cutoff estimate after EMA update as 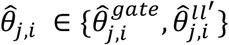 in the following sections.

##### 2.4.2.2 Response-adaptive Tailoring Functions

The second-stage randomization probabilities are determined by mapping the patient’s own stage-one outcome, *Y*_1*i*_, relative to the estimated cutoffs 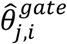 and 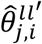. Let 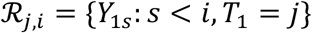 be defined as the reference set and denote the observed *Y*_1_s from the prior *i* − 1 patients who receive the same *T*_1_ = *j* as the current patient *i*. Let *y* ∈ ℛ_*j,i*_ be one of the observed *Y*_1_s. The following sections describe two RA-TF options.

###### 2.4.2.2.1 Percentile-Based Approach

This method utilizes percentile ranks to center the continuation probability around 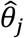. We compute the percentile rank of the current patient’s outcome *Y*_1*i*_ within ℛ_*j,i*_ as

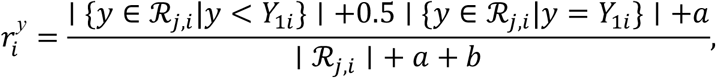

and the percentile rank of the estimated gate cutoff 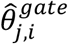 is computed as:

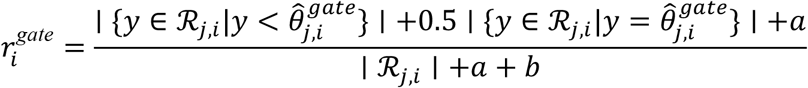

where 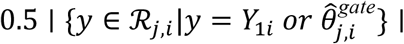 represents the mid-rank method to assign average rank to tied observations and it becomes 0 when no ties exist, and the additive constants *a* and *b* represent pseudo observations through Laplace smoothing^43^ to prevent extreme percentile estimates in the early stage. The continuous updating mechanism ensures that 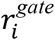 reflects the current best estimate of where the gate cutoff falls within the evolving outcome distribution for the *i*^*th*^ patient.

The corresponding continuation probability is defined as 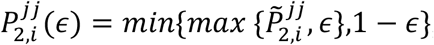, where 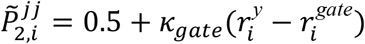. This centers the continuation probability at 0.5 when 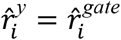 indicating that the percentile rank of the outcome for patient *i* is equal to the percentile rank of the estimated cut off, and adjusts proportionally based on the difference between 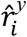 and 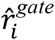 in percentiles (increase probability of staying on the same treatment when 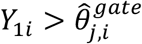, and decrease when for 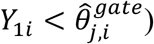. The sensitivity parameter *k*_*gate*_ controls how aggressively the randomization probabilities respond to the deviation between *Y*_1_ and the estimated 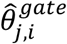. Values of *k*_*gate*_ > 1 amplify the difference between patient outcome and the cutoff, resulting in more extreme randomization probabilities for patients whose outcomes are distant from the cutoff. It produces stronger concentration of treatment allocation toward the optimal option and potentially accelerates convergence to superior treatments. However, this may lead to extremely unbalanced randomization across treatment sequences which reduces exploration and increases vulnerability to early misestimation of optimal thresholds. In contrast, values of *k*_*gate*_ < 1 mitigate the influence of outcome-threshold differences and maintain randomization probabilities closer to 0.5 across the outcome range to facilitate exploration of all treatment sequences. This conservative approach may require larger sample sizes to achieve substantial concentration on superior treatments but provides greater robustness against early estimation errors.

For patients who switch to alternative treatments, we compute the percentile rank of the estimated cutoff between two switching treatment options *l* and *l*′ ∈ ℒ_*j*_ (*l* ≠ *l*′ ≠ *j*) as

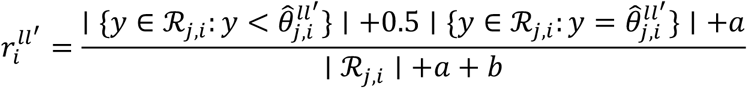

The conditional randomization probability of receiving treatment *l* among patients who switch from *j* is referred as 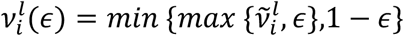,where 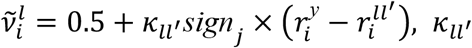 denotes the sensitivity parameter that controls the degree of adaptation between treatments *l* and *l*′ in parallel to *κ*_*gate*_ for the continuation decision; sign_*j*_ ∈ {−1, +1} captures the treatment effect orientation, which ensures patients with 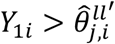 receive higher probability of switching to *l* when *l* is superior relative to *l*′ for higher outcomes (*sign*_*j*_ = +1). The conditional randomization probability of receiving treatment *l*′ among patients who switch from *j* is 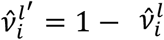.

##### 2.4.2.2.2 Standardized Score (Z-score) Approach

This method uses a modified *z*-score transformation centered at 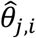, which accounts for the scale of variation in *Y*_1_. Based on the reference set ℛ_*j,i*_, we compute the mean and standard deviation of the observed *Y*_1_ within ℛ_*j,i*_

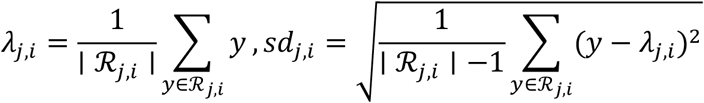

As in the percentile-based RA-TF, this approach also involves two sequential decisions. We first compute the z-score for current patient’s *Y*_1*i*_ centered at the estimated gate cutoff 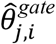 as

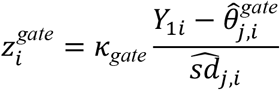

The continuation probability is then defined as 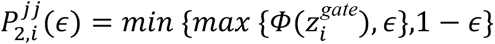, where Φ(·) denotes the standard normal cumulative distribution function. It centers the continuation probability at 0.5 when 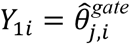, with continuation probabilities increasing for *Y*_1*i*_ above 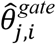 and decreasing for those below.

For patients who switch to alternative treatments, we compute the z-score of the estimated cutoff between *l* and *l*′ ∈ ℒ_*j*_ (*l* ≠ *l*′ ≠ *j*) as

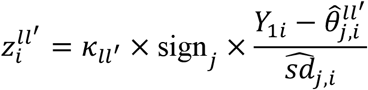

The conditional randomization probability of receiving treatment *l* among patients who switch from *j* is referred as 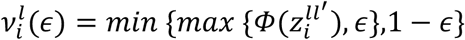. The conditional randomization probability of receiving treatment *l*′ among patients who switch from *j* is 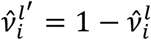.

For both RA-TFs, the overall second-stage randomization probabilities for patient *i* to switch to *l* and *l*′ ∈ ℒ, (*l* ≠ *l*′ ≠ *j*), are 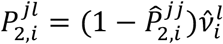 *and* 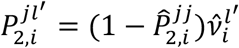, respectively.

## 3. DTR Effect Estimation Methods

The primary goal of DTR effect estimation is to identify the DTR which optimizes the expected second stage outcome, 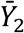, within a target population, termed the optimal DTR. This section focuses on outlining the weighting strategy and the application of Q-learning,^44-46^ tree-based reinforcement learning (TBRL)^47,48^ and G-estimation^49-51^ in the identification of optimal DTRs under BR-SMART, TF-SMART, GO-SMART and RA-TF-SMART frameworks. Covariates are excluded in the present study for simplicity, but all methods presented could be extended to incorporate covariates.

### 3.1 Weights

We employ IPTW to address potential selection bias arising from adaptive randomization and obtain unbiased estimates of DTR effects.^40,41^ Under the BR-SMART design, all participants are randomized with equal probability at each stage. Therefore, weighting is not required as the randomization structure itself ensures balance with *w*_*i*_(*T*_1*i*_, *T*_2*i*_) = 1 for all patients and all treatment sequences *d*(*T*_1*i*_, *T*_2*i*_).

In TF-SMARTs, all participants are randomized with equal probability in stage one and stage two between two switching treatments. Given the second-stage continuation probability defined as *f*(*Y*_1*i*_) for each patient *i*, the corresponding IPTW under our design is

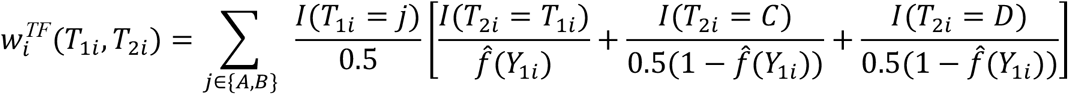

Under the GO-SMART and proposed RA-TF-SMART design, randomization probabilities are adaptive in both stages. During the burn-in period (*i* ≤ *n*_0_), patients are randomized as in BR-SMART with the same weight. For patients enrolled after the burn-in period (*i* > *n*_0_), weights incorporate both the response-adaptive first-stage probabilities and the tailoring function-based second-stage probabilities. The IPTW for GO-SMART is

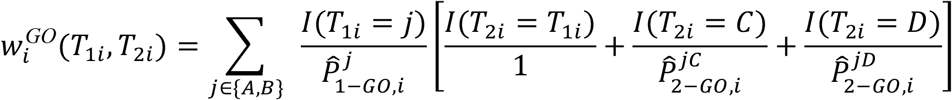

Similarly, the IPTW for RA-TF-SMART is

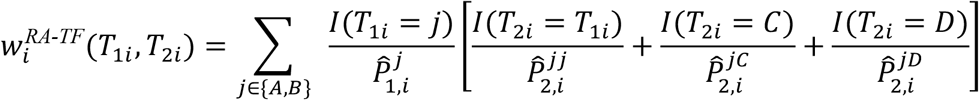

### 3.2 Q-learning

Q-learning is designed to maximize a continuous outcome to indirectly infer the optimal DTRs instead of directly optimizing the decision rule.^44-46^ In our two-stage SMART setting, Q-learning first defines the Q-function at the second stage as *Q*_2_(*T*_1_, *T*_2_, *Y*_1_) = *E*(*Y*_2_ ∣ *T*_1_, *T*_2_, *Y*_1_), which represents the expected second-stage outcome conditional on the treatment sequence and first-stage outcome. The optimal decision rule at the second stage is determined by identifying *T*_2_ that maximizes the second-stage Q-function as 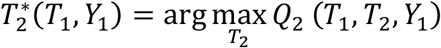.

To estimate and compare DTRs, linear regression-based Q-learning is implemented with IPTW applied to account for the randomization structure of the SMART design. The linear regression-based Q-function for the stage-two outcome is defined as

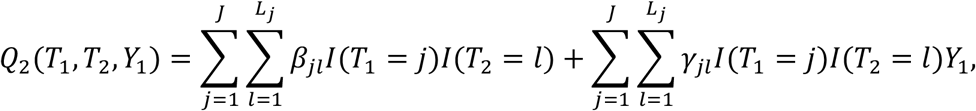

where *β*_*jl*_ is the intercept and *γ*_*jl*_ is the slope for the treatment sequences *d*(*j, l*). This model calculates the expected second-stage outcome, 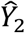, for all combinations of *T*_1_ and *T*_2_.

The method then proceeds recursively to stage one, where the Q-function incorporates a pseudo-outcome, 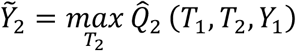, defined as the predicted second-stage counterfactual outcome under the optimal second-stage decision rule. The Q-function estimating the optimal first-stage decision rule based on 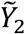 is defined as 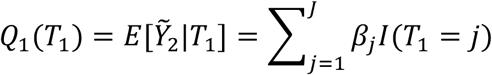, and the optimal *T*_1_ is defined as 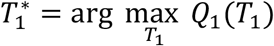, which is *T*_1_ = *j* with the largest 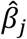.

### 3.3 Tree-based Reinforcement Learning

Prior studies have integrated reinforcement learning frameworks into decision tree algorithms to develop TBRL by adapting a tailored purity criterion designed for treatment decision-making.^47,48^ In TBRL, nodes represent subgroups of patients partitioned according to their characteristics, treatment history and intermediate outcomes, where the purity measure quantifies the benefit of assigning each subgroup to their corresponding treatment. We apply TBRL to build purity-based trees and identify optimal second-stage decision rules that maximize *Y*_2_, then select 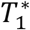 by comparing the second-stage pseudo-outcome, 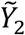.

As described in Laber and Zhao,^47^ the purity measure in the second stage is defined as:

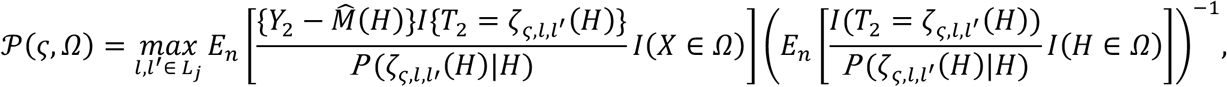

where Ω represents the node to be split into groups *ς* and *ς*^*c*^, *H* = {*Y*_1_, *T*_1_} represents patient intermediate outcome and treatment history, 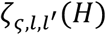 represents the decision rule that assigns treatment *l* to patients in group *ς* and treatment *l*′ to patients in group *ς*^*c*^ given 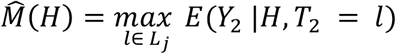 represents the expected outcome under optimal treatment given *H*, and *E*_*n*_ represents the empirical expectation operator. In the SMART setting, 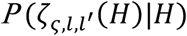 is captured by the IPTW. As a result, 𝒫 (*ς*, Ω) estimates the expected counterfactual *Y*_2_ for node Ω by the best decision rule that assigns *l* to all subjects in *ς* and *l*′ to all subjects in *ς*^*c*^.

We first perform recursive partitioning to grow the tree and find the optimal split in the second stage following the rules described in previous studies.^47,48^ The minimum node size and the minimum improvement in purity for a valid split is set as 10 and 0.00001, respectively. To determine the optimal split at each node, the observed outcomes are divided into percentiles, and each percentile value is evaluated as a candidate split point. The split that results in the greatest increase in the purity measure while fulfilling the node size requirement is identified. To estimate and compare the effects of DTRs, we adopt 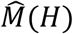 to model *Y*_2_ on *Y*_1_ and *T*_2_, and then define the pseudo-outcome as 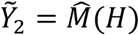. We use two methods for TBRL to estimate 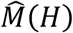. In the TBRL-LN method, the conditional mean model for continuous outcomes is mapped using a weighted linear regression similar to Q-learning. In contrast, TBRL-RF utilizes a non-parametric random forest algorithm to capture potentially complex and non-linear relationships. Then, the optimal *T*_1_ is identified as 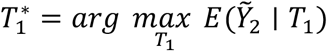, where 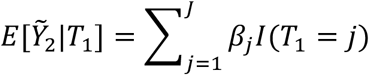.

### 3.4 G-Estimation

G-estimation is a semi-parametric method for estimating optimal DTRs under structural nested mean models (SNMM).^52^ We focus on a particular parameterization of the SNMM known as the optimal blip-to-zero function, which quantifies the expected benefit of receiving treatment versus a reference. The details of blip function theories and its general application form can be found in Chakraborty and Moodie. ^50^

For stage two, we set the blip function for continuing *T*_1_ = *j* as 0 and the linear blip function for switching to treatment *l* ∈ ℒ_*j*_ *where l* ≠ *j* is defined as 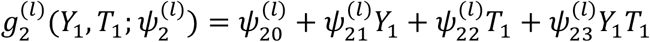, where 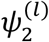 represents the set of blip function parameters 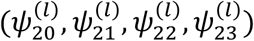. Positive blip values 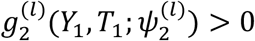 favor switching to option *l* over continuing *T*_1_. For stage 1, we specify an intercept-only blip function as *g*_1_(*ψ*_1_) = *ψ*_10_. G-estimation proceeds recursively from the final stage backward. At each stage, we construct pseudo-outcomes by removing the estimated effects of subsequent treatments under the optimal regime. For the second stage, we define the counterfactual outcome as 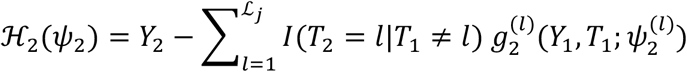, where *I*(*T* = *l* ∣ *T*_1_ ≠ *l*) represents the binary indicator for switching to option *l*. For notational convenience, we denote *I*_*l*_ ≡ *I*(*T*_2_ = *l* ∣ *T*_1_ ≠ *l*) . We construct instruments 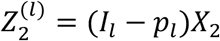 for each switch option *l*, where *X*_2_ = (1, *Y*_1_, *T*_1_, *Y*_1_*T*_1_)^⊤^ and *p*_*l*_ = *E*[*I*_*l*_ ∣ *Y*_1_, *T*_1_] represents the randomization probability which are known due to the SMART designs. These instruments are uncorrelated with ℋ_2_(*ψ*_2_) when *ψ*_2_ is equal to the true parameter values, 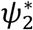. Let *Z*_2*i*_ represent the set of all instruments 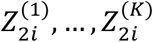 for patient *i* and *ψ*_2_ be the set of all blip parameters 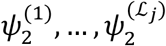, the doubly-robust estimating equation is defined as 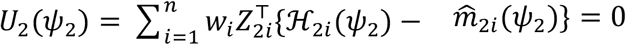, where *w*_*i*_ denotes the IPTW at stage 2 and 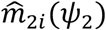 is an outcome regression obtained by regressing ℋ_2_(*ψ*_2_) on the basis *B*_2*i*_ = (1, *Y*_1*i*_, *T*_1*i*_, *Y*_1*i*_*T*_1*i*_) using weighted least squares. Then *ψ*_2_ is estimated by minimizing ∣∣ *U*_2_(*ψ*_2_) ∣∣^2^ . After obtaining 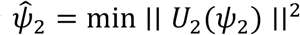, we construct the stage-1 pseudo-outcome as 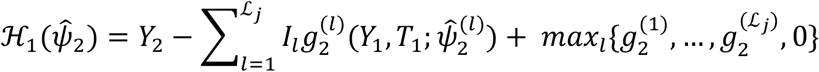, where 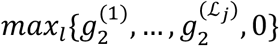 represents the expected benefit under the optimal stage-two decision. The stage-one estimating equation follows 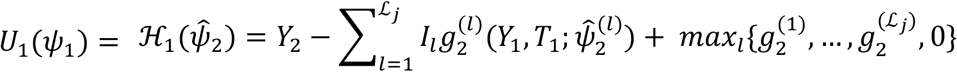, where 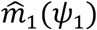 is fitted by weighted least squares on an intercept-only basis *B*_1*i*_ = (1) due to the absence of baseline covariates. The parameter *ψ*_10_ is estimated by minimizing ∣∣ *U*_1_(*ψ*_1_) ∣∣^2^.

The optimal treatment rules are determined by the estimated blip functions. At stage 1, treatment *T*_1_ = *A* is estimated to be optimal when 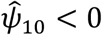. At stage 2, the optimal decision is to continue the first-stage treatment if 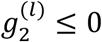 *for all l* ∈ ℒ_*j*_ *where l* ≠ *j*; otherwise, the switch option with the largest positive blip is selected. The stage-2 optimal rule is characterized by threshold values derived by solving 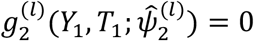 for *Y*_1_.

## 4. Simulation Study

In this section, we present simulation studies to evaluate the performance of the RA-TF-SMART designs across the two proposed RA-TFs as outlined in **Sections 2.4**, and compare against BR-SMART, TF-SMART, and GO-SMART using the analytical methods introduced in **Section 3**. We propose nine scenarios with diverse treatment effect parameters to represent various clinical decision-making patterns that may be encountered when developing DTRs (**Table 1, Supplementary Table 1**). For each scenario under the various SMART designs, we simulate *n* = 300 patients for each trial and replicate the simulation 1,000 times.

**Table 1.**
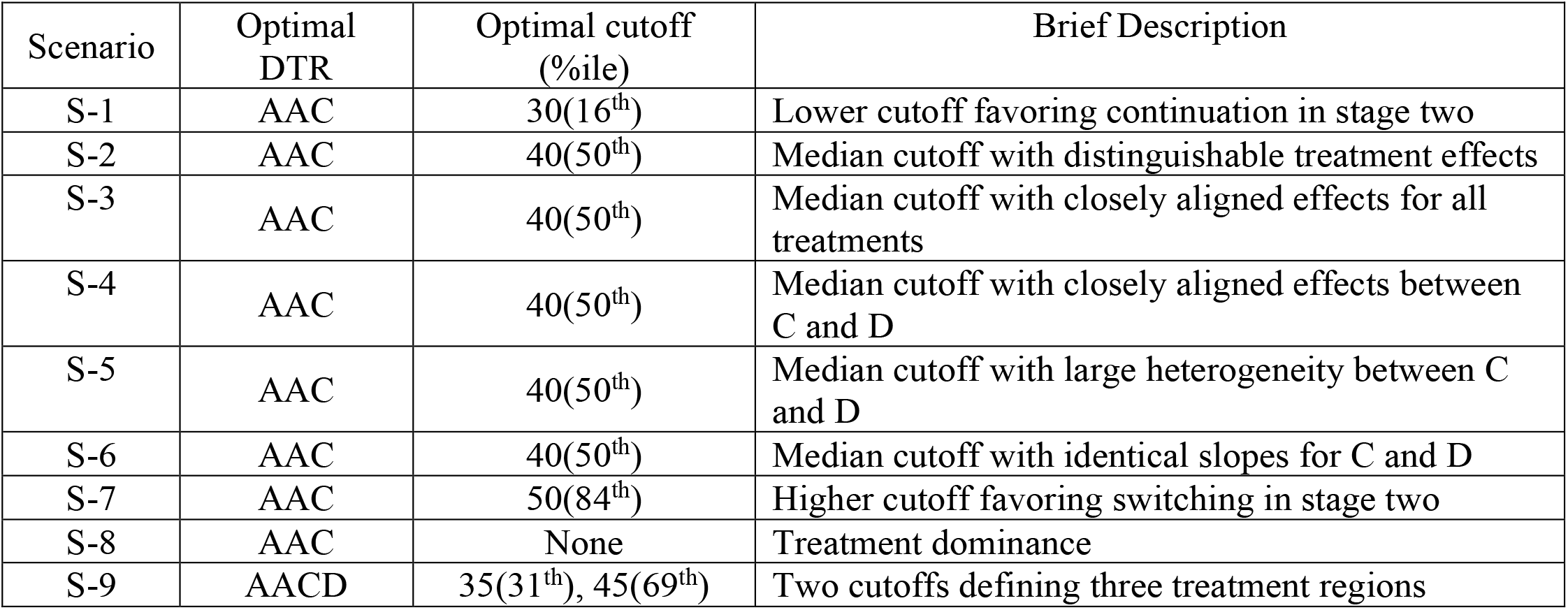
Summary of simulation scenarios with theoretically optimal dynamic treatment regimens and decision cutoffs.

### 4.1 Data-Generating Mechanism

After first-stage randomization according to the SMART designs, the stage 1 outcome *Y*_1*i*_ for patient *i* assigned to treatment *j* is generated based on a treatment-specific normal distribution: 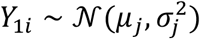 where *μ*_*j*_ represents the mean first-stage outcome and 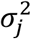 represents the variance for first-stage treatment *j*. The second-stage treatment *l* is assigned according to the randomization probabilities determined by the different SMART designs described in **Sections 2**. According to our assumption in **Section 2.1**, the expected value of *Y*_2*i*_ is defined as *μ*(*T*_1*i*_ = *j, T*_2*i*_ = *l, Y*_1*i*_) = *β*_*jl*_ + *γ*_*jl*_*Y*_1*i*_, where *β*_*jl*_ represents the treatment-sequence-specific intercept and *γ*_*jl*_ represents the treatment-sequence-specific slope coefficient. The second stage outcome is then generated from a patient-specific distribution defined as: 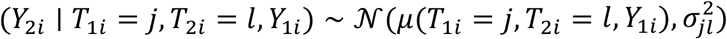, where 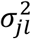 represents the variance for treatment sequence *d*(*j, l*).

In this study, we include two first-stage treatment options: A and B. We assume treatment A is superior in the first stage with *μ*_*A*_ = 40 and *μ*_*B*_ = 30, and set a common standard deviation of *σ* = 10 for both stage-one treatments. The second-stage outcomes are generated by varying the intercept *β*_*jl*_ and slope *γ*_*jl*_ according to the scenario-specific settings presented in **Supplementary Table 1**, and a common standard deviation of *σ* = 10 across treatment sequences.

### 4.2 Theoretical Optimal DTR and Cutoffs Determination

For each scenario, we determine the theoretically optimal DTR and corresponding cutoffs based on the data-generating parameters specified in **Supplementary Table 1**. These theoretical values serve as benchmarks for evaluating the performance of different SMART designs and estimation methods.

#### 4.2.1 Optimal DTR Identification

The theoretical optimal DTR for each patient is defined as the treatment sequence that maximizes the expected *Y*_2_ given their *Y*_1_. Using the data-generating model from **Section 4.1**, the optimal DTR is determined by comparing the expected second-stage outcomes across all available treatment sequences:

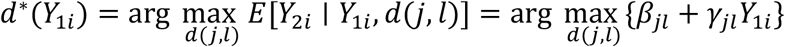

where *d*(*j, l*) represents a treatment sequence consisting of *j* ∈ 𝒥 and *l* ∈ ℒ_*j*_. This optimal DTR varies across patients based on their individual *Y*_1*i*_ values and is determined by the intersection points of the linear functions defined by the treatment-sequence-specific parameters. It is important to note that the theoretical optimal DTR for each patient is not observable in practice, as we cannot observe the full set of counterfactual outcomes under all possible treatment sequences for any given patient.

#### 4.2.2 Optimal Cutoff Calculation

As described in **Section 2.4.2.1**, the optimal second-stage cutoff value *θ*^*^ represents the *Y*_1_ value at which the expected second-stage outcomes, 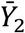, under two *T*_2_ options are equal. Specifically, since we set treatment A as the optimal first-stage treatment in all scenarios, this cutoff is calculated by solving the equation 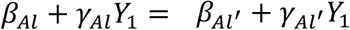, where 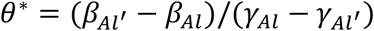.

Our design with three second-stage options (continue with A, switch to C, or switch to D) may produce a single cutoff representing one boundary separating two treatment recommendations across the *Y*_1_ range, two cutoffs defining three regions where each region favors a different second-stage treatment, or no cutoff when one treatment dominates across all *Y*_1_ values or all treatments yield equivalent outcomes.

### 4.3 Simulation Parameters

#### 4.3.1 Scenario Specifications

**Table 1** displays the optimal DTR and optimal cutoffs for second-stage treatment decisions among patients who received the optimal first-stage treatment A across nine scenarios we propose. S-1 through S-7 use a single cutoff design where treatment C consistently outperforms treatment D across the *Y*_1_ range. Under these scenarios, the optimal second-stage treatment is A when *Y*_1_ exceeds the cutoff value and C when *Y*_1_ falls below the cutoff. We vary the parameters across S-1 through S-7 to examine the influence of cutoff value and treatment effect difference on the performance of SMART designs. Specifically, S-1 establishes a lower cutoff at 30 for around 16^th^ percentile of the *Y*_1_ distribution to favor continuation in the second stage. S-2 through S-6 all employ a cutoff of 40 equal to the *Y*_1_ mean (50^th^ percentile) to balance continuation and switching in the second stage but different design parameters to isolate specific performance factors. S-2 serves as the basic scenario with reasonably distinguishable treatment coefficient difference across three *T*_2_ options. S-3 specifies the treatment effect of *T*_1_ aligned closely to two switching treatments (C and D). S-4 keeps distinguishable treatment effect between *T*_1_ and the optimal switching treatment C, while creating closely aligned treatment effect coefficients for switching treatments C and D. S-5 also maintains distinguishable treatment effect between the first-stage treatment A and the optimal switching treatment C, while introducing substantially larger heterogeneity between switching treatments C and D. S-6 sets the treatment slope for treatment C and D to be identical to eliminate the predictive capability of *Y*_1_ for *Y*_2_. S-7 establishes a higher cutoff of 50 for around 84^th^ percentile of the *Y*_1_ distribution to favor switching in the second stage. S-8 examines treatment dominance without cutoff for *T*_2_ decisions such that continuation of treatment A consistently outperforms all alternatives regardless of *Y*_1_ values. Scenario S-9 establishes two cutoffs defining three response regions for optimal second-stage treatment assignment, where the cutoff between continuation and switching is 45 for around 69^th^ percentile of the *Y*_1_ distribution. In this scenario, we denote the optimal DTR as AACD to indicate that when *Y*_1_ exceeds the second cutoff, A is the optimal treatment; when *Y*_1_ falls between the first and second cutoffs, C is optimal; and when *Y*_1_ falls below the first cutoff, D is the optimal treatment.

#### 4.3.2 SMART Design Parameters

For TF-SMART designs, we consider three TFs modified by the adaptation intensity parameter *τ* ∈ {0.5,1,2}: (1) TF1: *f*(*Y*_1*i*_) = *Y*_1*i*_/100; (2) TF1/2: *f*(*Y*_1*i*_) = (*Y*_1*i*_/100)^1/2^; and (3) TF2: *f*(*Y*_1*i*_) = (*Y*_1*i*_/100)^2^.

In the GO-SMART designs, a burn-in period comprising 20% of the total sample *n* = 300 (*n*_0_ = 60) is used to initialize the adaptive randomization process. We also set the adaptation intensity parameter *τ* = 1 and the constraint parameter *ϵ* = 0.1. Additionally, we set a conservative second-stage response threshold *φ*_2_ = 40 for all DTRs based on our outcome simulation parameters to avoid invalid response rates across all treatment sequences. We further examine the stage-one response threshold across *φ*_1_ ∈ {20,30,40,50,60}, denoted as GO-SMART 20 through GO-SMART 60, to evaluate the impact of the first-stage cutoff dichotomization on performance.

For RA-TF-SMART designs, and in the GO-SMART designs, we set the burn-in proportion *p*_0_ = 0.2, the adaptation intensity parameter *κ*_*gate*_ = *κ*_*CD*_ = 1, and the constraint parameter *ϵ* = 0.1. For the second-stage RA-TFs, we set the EMA smoothing parameter *η* = 0.9 to assign 90% weight to the accumulated historical estimate and 10% weight to the new estimate, as well as moderate Laplace smoothing parameters *a* = 5, *b* = 5.

### 4.4. Performance Evaluation

#### 4.4.1 Comparison of Trial-Level Metrics

We first evaluate the trial-level outcomes to compare the performance of SMART designs. For the RATF-SMART and GO-SMART designs, we remove the 60 burn-in patients to focus on the adaptive portion of the trial. For the BR-SMART and TF-SMART, we randomly remove 60 patients to maintain comparable effective sample sizes. We calculate the proportion of patients randomized to the superior first-stage treatment A in the trial, which evaluates whether the RAR successfully shifts randomization toward the favorable treatment. Different SMART designs incorporate different first-stage randomization strategies. The varying proportions of patients assigned to the superior first-stage treatment A impacts overall trial performance. To enable fair comparisons, we focus our analysis on patients who receive treatment A in the first stage. For this subgroup, we first compute the probability of continuing with treatment A in the second stage, denoted as *P*_*AA*_. This metric reflects how the RA-TFs respond to the distribution of first-stage outcomes and the location of the theoretical optimal cutoff. We then calculate the mean second-stage trial outcome, 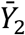, which reflects the overall patient benefit during the trial itself. Finally, we determine the proportion of patients whose second-stage treatment assignment matches the theoretically optimal treatment according to the true DTR, denoted as 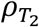. This metric evaluates the alignment between RA-TF treatment assignments and optimal treatment decisions throughout the trial.

#### 4.4.2 Comparison of DTR estimation

To evaluate the performance of DTR estimation, we apply each estimated optimal DTR to an independent validation sample of 10,000 patients generated from the same data-generating mechanism. We estimate the optimal DTR using Q-learning, TBRL-LN, TBRL-RF, and G-estimation from the simulated trial data as described in **Sections 4.1** and **4.2**, then assign treatments to the validation sample according to each estimated DTR and calculate the following metrics.

##### 4.4.2.1 Across Designs and Scenarios

Across all SMART designs, we first calculate the proportion of simulation replications that fail to produce cutoff estimates, 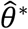, consistent with the scenario structures. In S-1 through S-7 with a single cutoff, undefined cutoffs 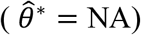 represent estimation failures. A high proportion of undefined cutoffs indicates correct estimation in S-8 and the identification of two cutoffs represent successful estimation in S-9. For the rest metrics, we restrict the comparison to replications that correctly identify the scenario structure: one cutoff for S-1 through S-7, no cutoff for S-8, and two cutoffs for S-9. We compare 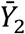 and 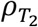 to examine whether incorporating RAR compromises post-trial DTR estimation accuracy relative to other SMART designs. Additionally, we compare the 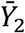 obtained from the estimated DTRs to the theoretical 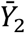 obtained from applying the theoretically optimal DTRs to the same validation population. We quantify the accuracy of cutoff estimation through bias defined as the difference between the estimated cutoff and the true optimal cutoff described in **Table 1**. We also estimate the proportion of simulation replications that correctly identify treatment A as the optimal first-stage treatment, 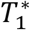.

##### 4.4.2.2 Across Analytical Methods

Within each SMART design and scenario, we compare the four analytical methods (Q-learning, TBRL-LN, TBRL-RF, G-estimation) across the same metrics described in **Sections 4.4.2.1**, including the proportion of simulation replications with 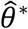 consistent with the scenario structures, 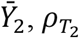, cutoff bias, and the proportion of simulation replications that correctly identify treatment A as 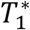.

## 5. Results

### 5.1 Trial-Level Metrics

The comparison of trial-level metrics reveals a consistent pattern favoring RA-TF-SMART designs over other adaptive SMARTs and BR-SMART. The proportion of patients randomized to the superior first-stage treatment A depends exclusively on the randomization mechanism of each SMART since all scenarios share identical stage-one outcome distributions (**Supplementary Figure 1**). BR-SMART and TF-SMART maintain balanced allocation (*P*(*T*_1_ = *A*) = 0.50), whereas RA-TF-SMART introduces a moderate adaptive shift toward A (*P*(*T*_1_ = *A*) = 0.5_7_). In contrast, GO-SMART shows more aggressive allocation toward Treatment A, reaching the maximum *P*(*T*_1_ = *A*) = 0.85 when the first-stage response cutoff is set at 50. This pattern reflects the sensitivity of GO-SMART to the dichotomization rules where both extremely low (e.g., 20) or high (e.g., 60) cutoffs compress the response rates of both first-stage treatments and weaken adaptation, while intermediate cutoffs (e.g., 30–50) amplifies differences in binary response proportions and drive aggressive allocation toward A.

Only RA-TF-SMART consistently responds to treatment effect heterogeneity and generates appropriate *P*_*AA*_ (*P*(*T*_2_ = *A*|*T*_1_ = *A*)) in the second stage across scenarios through adaptive randomization (**Figure 2**). This adaptive property stems from the centering process achieved through recursive estimation of the optimal continuation cutoff as outcome data accumulates. Specifically, RA-TF-SMART correctly favors continuation for approximately 72% of patients in S-1 and 25-27% in S-7. For S-2 through S-6, RA-TF-SMART maintains *P*_*AA*_ close to 50%, except in S-3 where the treatment effect between continuation and switching is closely aligned. In contrast, the continuation probabilities of the other SMART designs remain constant across scenarios since they do not incorporate treatment effect estimation to guide second-stage randomization. BR-SMART preserves balanced randomization probabilities (1/*L*_*A*_ = 0.33) regardless of observed outcomes. TF-SMART applies a predetermined linear transformation based on the measurement scale of *Y*_1_ without adaptation for the observed *Y*_1_ distribution. As a result, TF 1 mistakenly assigns 40% of patients to continuation in S-2 through S-6 where around 50% of the population benefits from remaining on treatment A. In addition, GO-SMART determines continuation exclusively through a fixed response cutoff and assigns all first stage “responders” to continuation. When the GO-SMART response cutoff coincides with the scenario’s true cutoff, GO-SMART assigns the correct proportion of patients to continuation. However, RA-TF-SMART generates continuation probabilities that are slightly more conservative due to its randomization mechanism intentionally retaining non-zero probability for alternative treatments among patients with favorable outcomes (i.e., 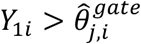).

**Figure 2.**
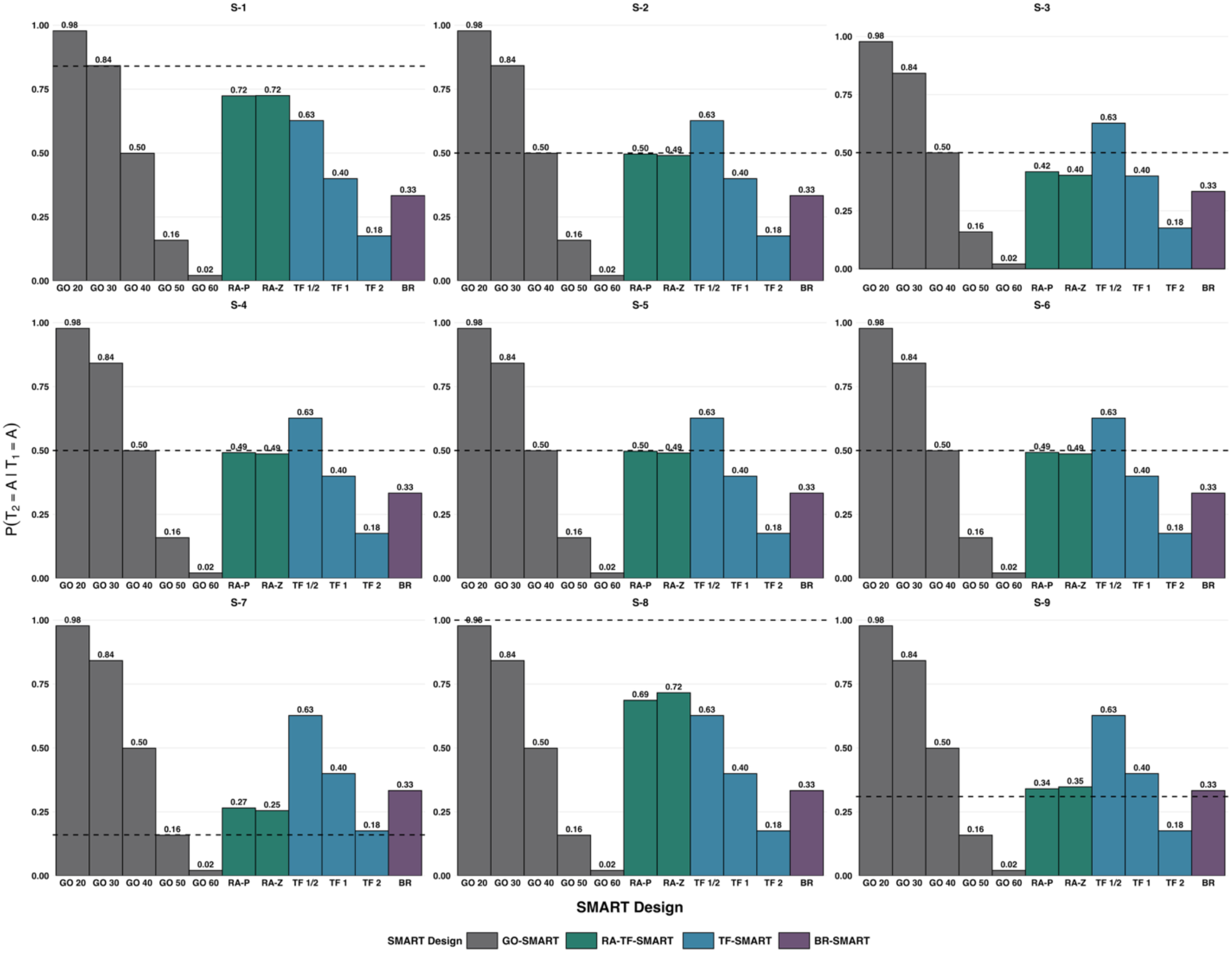
Probability of continuing treatment A in stage two among patients who received treatment A in stage one across sequential multiple assignment randomized trial (SMART) designs and simulation scenarios. Dashed lines indicate the theoretical optimal probability of continuation under each scenario. Abbreviations: GO 20 through GO 60, generalized outcome-adaptive SMART with stage-one response cutoffs of 20 to 60; RA-P, percentile-based response-adaptive tailoring function SMART; RA-Z, Z-score response-adaptive tailoring function SMART; TF-½, TF-1, and TF-2, tailoring function SMART with adaptation intensity parameters of ½, 1, and 2; BR, balanced randomization SMART.

**Figures 3** and **4** reveal that the BR-SMART and TF-SMARTs consistently generate weak trial outcomes due to the lack of between-patient adaptation, while RA-TF-SMART designs achieve 20-40 absolute percentage point improvements in 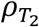 (the proportion of patients whose second-stage treatment assignment matches the theoretically optimal treatment according to the true DTR) and consistently higher 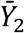. The z-score RA-TF slightly outperforms percentile-based RA-TF. GO-SMARTs show competitive performance to RA-TF-SMARTs but are highly dependent on the first-stage response cutoff and treatment effect heterogeneity. Several scenarios demonstrate notable advantages of RA-TF-SMARTs over GO-SMARTs. In S-7, RA-TF-SMART designs achieve higher 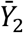 (73.16-74.53) than GO-SMART across all cutoff values (68.55-72.55). This pattern supports the advantage of RA-TF-SMART to respond treatment effect heterogeneity and generate appropriate second-stage randomization probability favoring switching. GO-SMARTs outperform RA-TF-SMART when the cutoff selection aligns closely with the scenario structure and generate higher 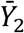 and 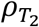, especially in scenarios favoring continuation. For example, GO-SMART 20 achieves 26% higher 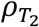 and dramatically higher outcomes of 87.56 than RA-TF-SMART (79.27-80.06) in S-8 by maximizing continuation assignments in a scenario where continuation is dominant across *Y*_1_ range. This advantage derives from GO-SMART’s aggressive randomization strategy to assigns all responders to continue with *T*_1_ and maximizes the trial benefit, whereas RA-TF-SMART retains a percentage of patients receiving alternative treatments even when continuation is superior. This property also reflects an important limitation of GO-SMART that higher 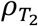 does not always guarantee better 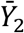. In S-1, S-2, S-3 and S-6, the cutoff resulting in the highest 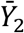 always tends to be lower than the theoretical cutoff that generate the highest 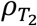. For example, in S-2, GO-SMART 30 achieves the highest 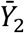 of 69.42 despite the theoretical optimal cutoff of 40 assigns the highest 83% patients to their optimal *T*_2_. This discrepancy stems from GO-SMART’s inability to effectively distinguish between the two alternative treatments C and D when using the arbitrary second-stage response cutoff of 40. That is, both C and D lead to similar response rates due to the dichotomization of the continuous outcome to define response. In these scenarios, there are more non-responders who are then randomized between C and D in GO-SMART 40 compared to GO-SMART 30, which leads to more misallocation that reduces 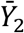. This limitation is mitigated in S-4 and S-5, where the cutoff of 40 generates the highest outcomes because the similarity between the effects of C and D reduces the penalty of inaccurate second-stage allocation (S-4) or the extreme treatment differences force GO-SMART to better distinguish C and D (S-5). In addition, in S-3 where treatment A is closely aligned with both C and D and S-9 with two decision cutoffs, the 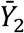 differences among all designs are relatively minimal. This indicates that RA-TF-SMARTs and GO-SMARTs all struggle to detect ambiguous treatment differentiation or guide complex decision rules. However, RA-TF-SMARTs still successfully improves 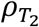 from 0.33 in BR-SMART to over 0.5.

**Figure 3.**
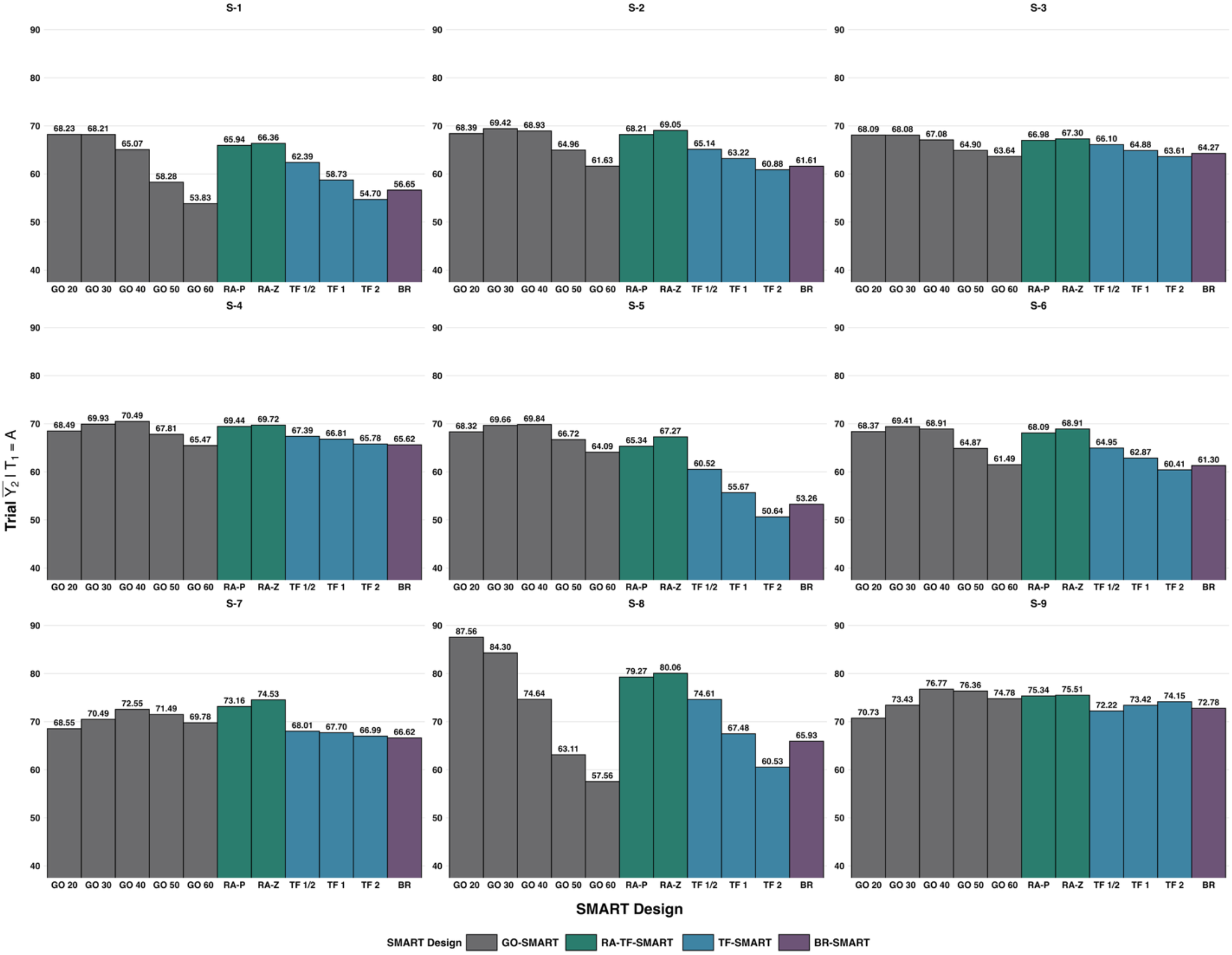
Average second-stage outcome among patients who received treatment A in stage one across scenarios and sequential multiple assignment randomized trial (SMART) designs. Abbreviations: GO 20 through GO 60, generalized outcome-adaptive SMART with stage-one response cutoffs of 20 to 60; RA-P, percentile-based response-adaptive tailoring function SMART; RA-Z, Z-score response-adaptive tailoring function SMART; TF-½, TF-1, and TF-2, tailoring function SMART with adaptation intensity parameters of ½, 1, and 2; BR, balanced randomization SMART.

**Figure 4.**
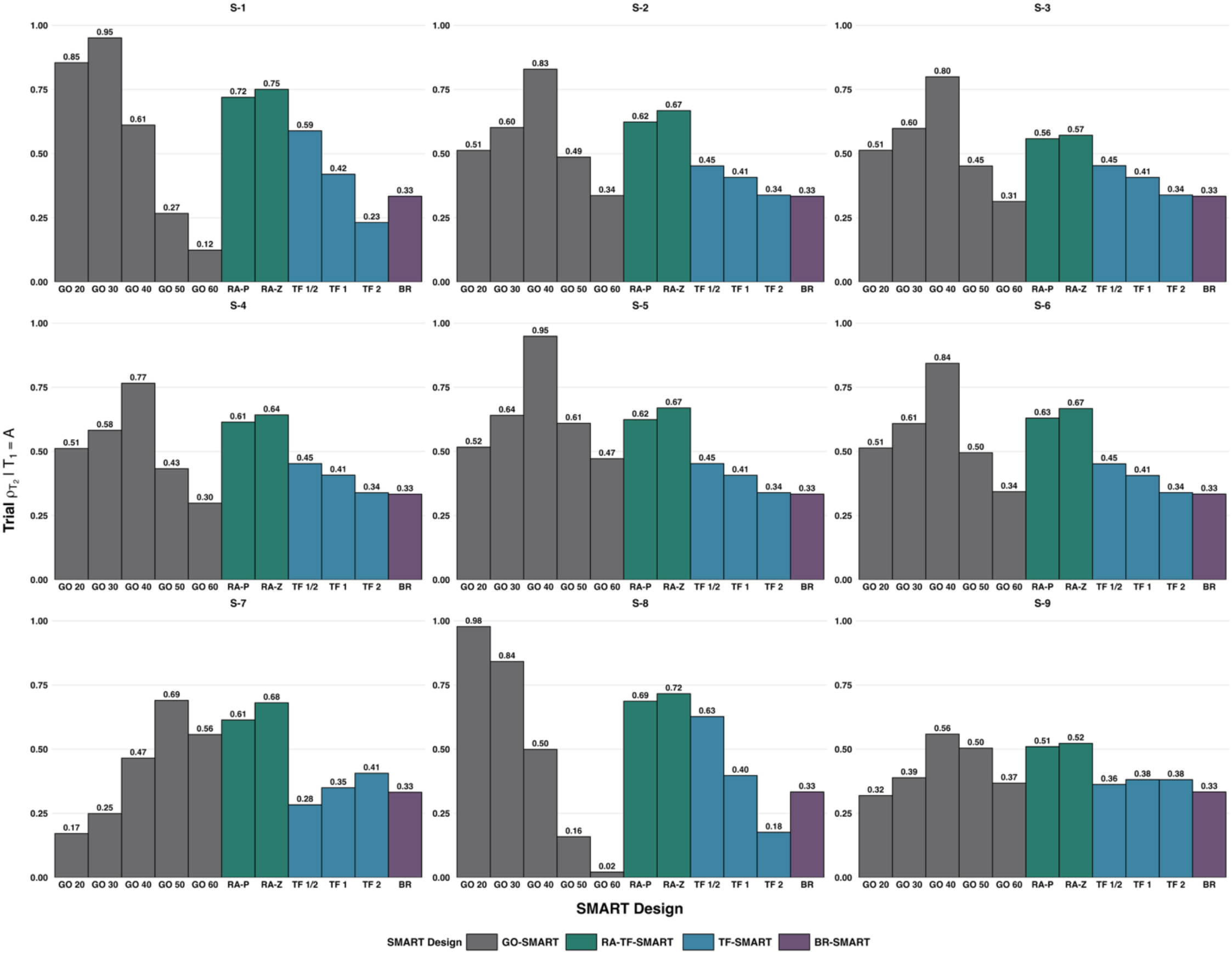
Proportion of second-stage randomization following the optimal dynamic treatment regimen among patients who received treatment A in stage one across sequential multiple assignment randomized trial (SMART) designs and scenarios. Abbreviations: GO 20 through GO 60, generalized outcome-adaptive SMART with stage-one response cutoffs of 20 to 60; RA-P, percentile-based response-adaptive tailoring function SMART; RA-Z, Z-score response-adaptive tailoring function SMART; TF-½, TF-1, and TF-2, tailoring function SMART with adaptation intensity parameters of ½, 1, and 2; BR, balanced randomization SMART.

### 5.2 Estimates by Scenarios and SMART Designs

As shown in **Supplementary Figure 2**, different SMART designs generally produce similar invalid cutoff estimate 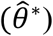 proportions although GO-SMART demonstrates more fluctuations across response cutoffs. This indicates that the RA-TF-SMART design does not compromise estimation reliability compared to BR-SMART. Across nine scenarios, S-3 with minimal treatment differentiation and S-9 with two decision cutoffs demonstrate higher invalid proportions across designs and methods. In addition, S-1 and S-7 with cutoffs at distributional extremes show slightly elevated invalid proportions. This indicates that weak treatment differentiation, decision complexity, and limited data near the decision cutoff region directly impair cutoff estimation. Furthermore, Scenario 6 supports that the removal of predictive probability of *Y*_1_ does not compromise DTR estimation.

**Figures 5 and 6** display the distribution of 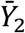 and 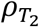 across 1,000 simulation replications when applying estimated DTRs to a validation population of patients. Overall, BR-SMART provides the most robust performance across all scenarios, with tightly clustered 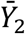 distributions whose medians best approximate theoretical reference values and the highest median 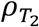. RA-TF-SMARTs consistently produce comparable 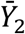 and 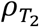 distributions to BR-SMART but with slightly greater variability. The performance of GO-SMART is generally worse than RA-TF-SMART with lower median 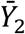 and 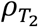. GO-SMART improves when the dichotomization cutoff tends to align with *θ*^*^ (e.g., GO-30 in S-1 and GO-40 in S-2). In addition, the estimation of GO-SMART designs shows greater variability and more outliers, especially when the response cutoff is extreme (e.g., GO-20 or GO-60). **Figure 7** also confirms that the estimation of *θ*^*^ in GO-SMARTs is less stable with more outliers compared to RA-TF-SMARTs. Between the two RA-TFs, the percentile-based method generates a lower proportion of invalid *θ*^*^ and more stable *θ*^*^ estimates.

**Figure 5.**
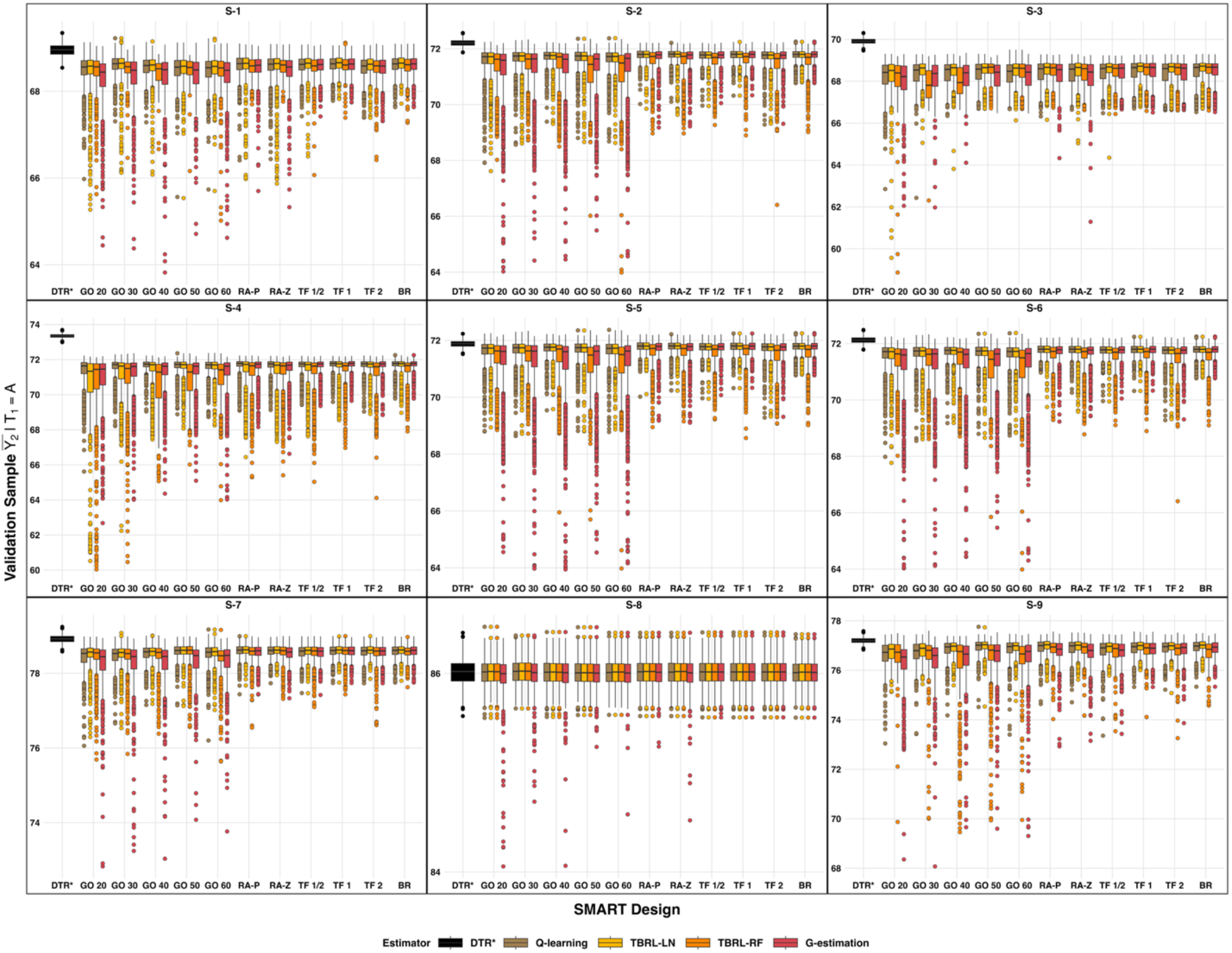
Average second-stage outcome when applying estimated dynamic treatment regimens in the validation sample across simulation scenarios, sequential multiple assignment randomized trial (SMART) designs, and analytical methods. Abbreviations: DTR*, optimal dynamic treatment regimen; TBRL-LN = tree-based reinforcement learning with linear regression; TBRL-RF = tree-based reinforcement learning with random forest; GO 20 through GO 60, generalized outcome-adaptive SMART with stage-one response cutoffs of 20 to 60; RA-P, percentile-based response-adaptive tailoring function SMART; RA-Z, Z-score response-adaptive tailoring function SMART; TF-½, TF-1, and TF-2, tailoring function SMART with adaptation intensity parameters of ½, 1, and 2; BR, balanced randomization SMART.

**Figure 6.**
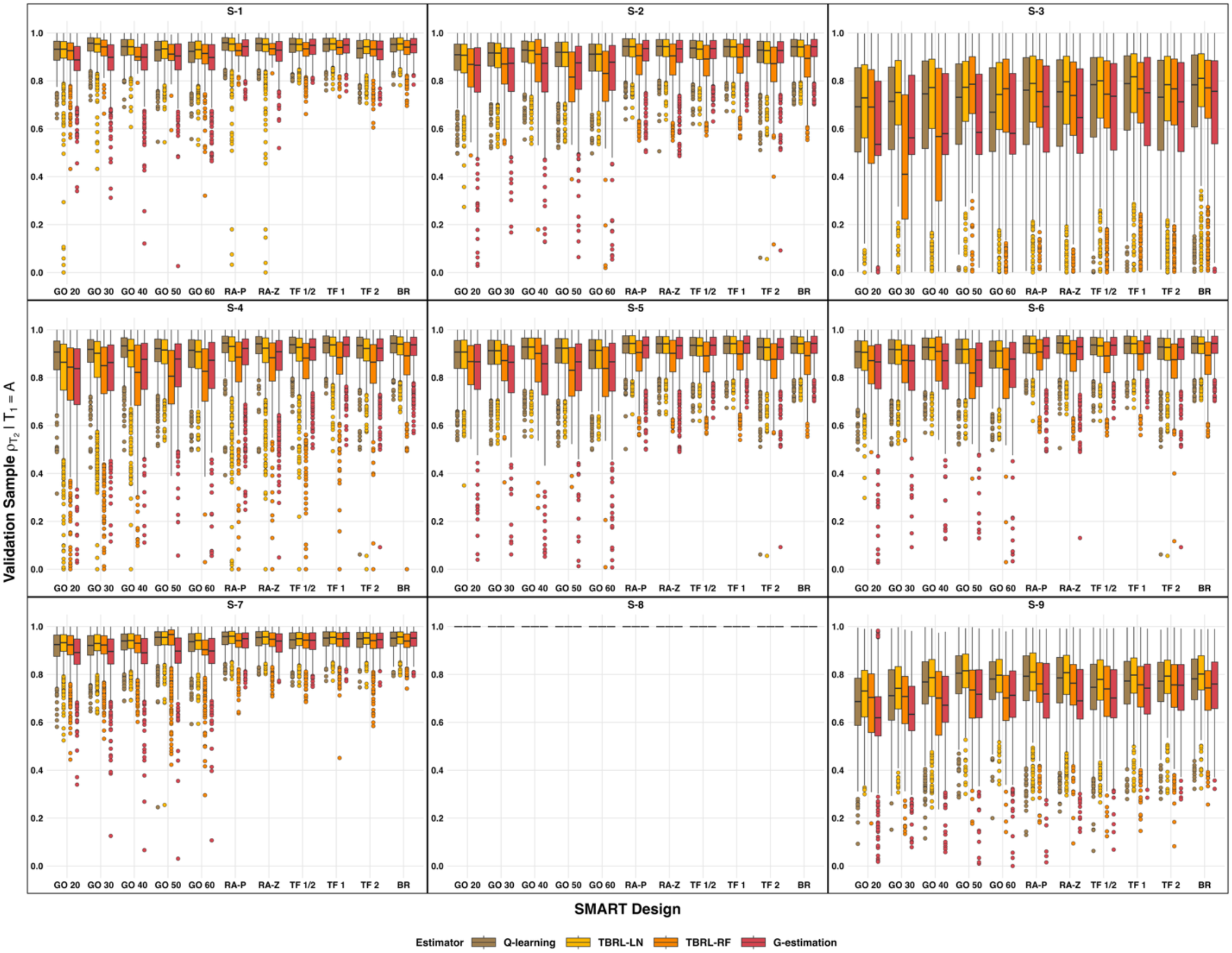
Proportion of second-stage randomization when applying estimated dynamic treatment regimens in the validation sample across simulation scenarios, sequential multiple assignment randomized trial (SMART) designs, and analytical methods. Abbreviations: DTR, dynamic treatment regimen; TBRL-LN = tree-based reinforcement learning with linear regression; TBRL-RF = tree-based reinforcement learning with random forest; GO 20 through GO 60, generalized outcome-adaptive SMART with stage-one response cutoffs of 20 to 60; RA-P, percentile-based response-adaptive tailoring function SMART; RA-Z, Z-score response-adaptive tailoring function SMART; TF-½, TF-1, and TF-2, tailoring function SMART with adaptation intensity parameters of ½, 1, and 2; BR, balanced randomization SMART.

**Figure 7.**
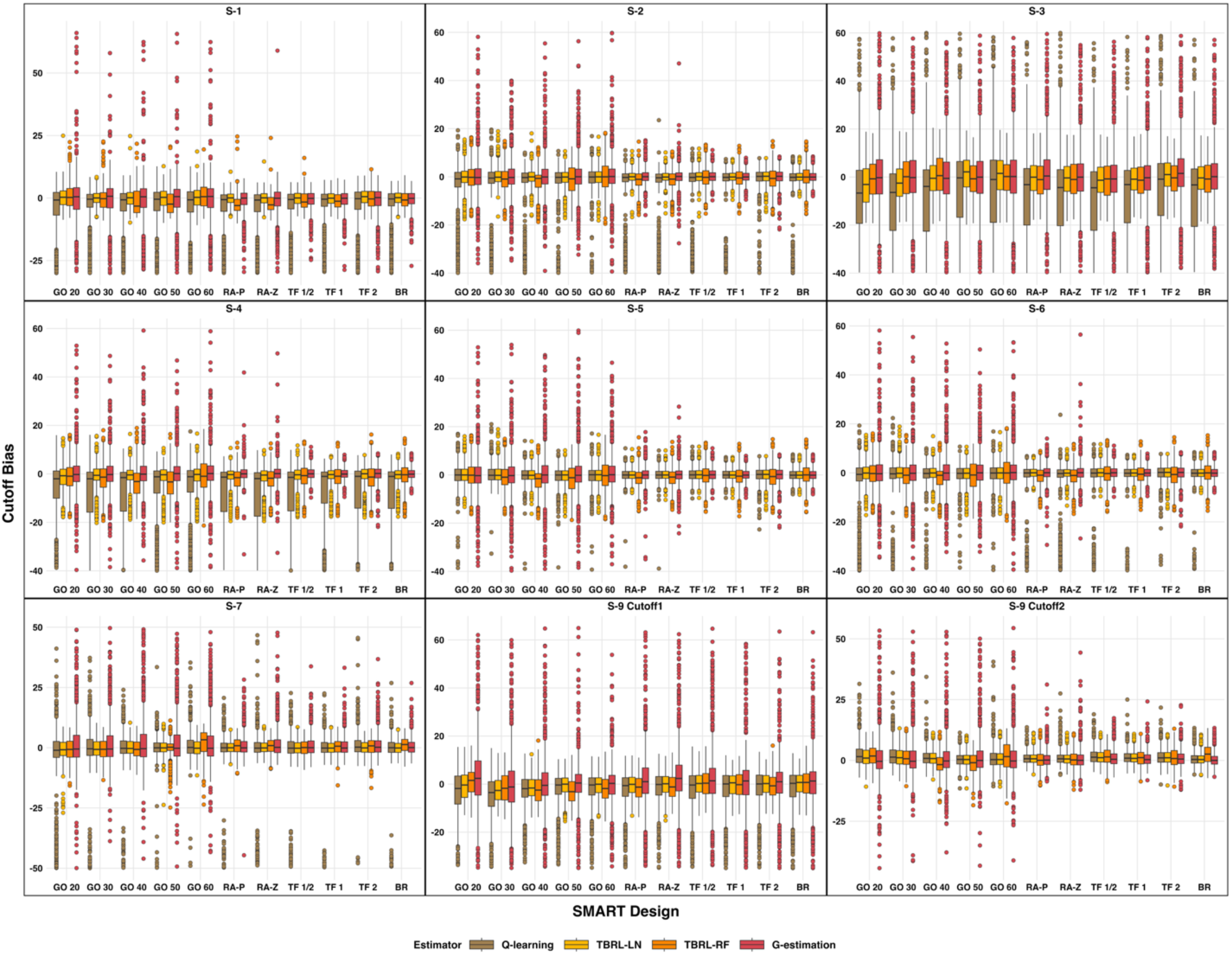
Estimated cutoff bias relative to the optimal cut off when applying estimated dynamic treatment regimens in the validation sample across simulation scenarios, sequential multiple assignment randomized trial (SMART) designs, and analytical methods. Abbreviations: TBRL-LN = tree-based reinforcement learning with linear regression; TBRL-RF = tree-based reinforcement learning with random forest; GO 20 through GO 60, generalized outcome-adaptive SMART with stage-one response cutoffs of 20 to 60; RA-P, percentile-based response-adaptive tailoring function SMART; RA-Z, Z-score response-adaptive tailoring function SMART; TF-½, TF-1, and TF-2, tailoring function SMART with adaptation intensity parameters of ½, 1, and 2; BR, balanced randomization SMART.

Across scenarios, the performance shifts according to the underlying treatment effect structure (**Figure 5-6**). In S-2, S-4, and S-5 where the distinction between continuation and switching is clear, all SMART designs produce 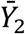 distributions close to the optimal reference and median 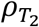 higher than 0.8. In contrast, in S-3 and S-4, we find the median estimated 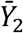 is much lower than the optimal value with higher variability and the median 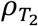 is below 0.8 in S-3. Additionally, S-1 and S-7 have to higher treatment assignment accuracy than S-2 through S-6 due to the distributional properties of the RA-TFs and their interaction with *θ*^*^ location. When *θ*^*^ is located around the distribution median, a substantial proportion of participants have *Y*_1_ clustered near the center of the distribution with corresponding continuation probabilities around 0.50. As a result, participants retain relatively high probability of being randomized to alternative treatments even when continuation represents the optimal treatment. In contrast, when *θ*^*^ falls at the 25th or 75th percentile, the majority of participants have *Y*_1_ clearly above or below *θ*^*^, resulting in more decisive randomization continuation/switching probabilities such as 0.70 to 0.90. The estimation in S-8 with treatment dominance is consistently good across SMART designs since we focus on replications without identifying valid *θ*^*^. Under the two-cutoff setting in S-9, all designs experience a reduction in 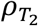 relative to single-cutoff scenarios, but RA-TF-SMARTs remain the most robust among the adaptive approaches with median 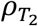 around 0.8. Additionally, treatment A is correctly identified as the optimal stage one treatment, 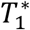, for all SMART designs across each scenario.

### 5.3 Estimates by Analytical Methods

Across all metrics, Q-learning consistently demonstrates the most reliable performance, with median 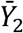 closer to the optimal DTR, higher median 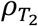 around 0.9, and precise cutoff estimates with median bias near zero (**Figures 5-7**). In addition, Q-learning generally maintains the lowest invalid *θ*^*^ proportions to ensure nearly all replications contribute to DTR identification. This reflects the advantage of parametric regression models with IPTW when the assumed linear relationship holds. TBRL-LN shows comparable performance to Q-learning for 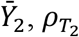, and *θ*^*^ estimation but produces higher invalid *θ*^*^proportions. TBRL-LN demonstrates concentrated 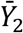 and 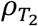 distributions with fewer outliers than Q-learning in S-3 and S-9, which may be attributed to higher percentages of replications with invalid cutoffs that we remove from the analyses. Both Q-learning and TBRL-LN struggle with *θ*^*^ estimation in S-3 and S-4 where the treatment effect is minimal. G-estimation shows competitive performance in estimating 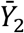 and cutoffs that are generally consistent with Q-learning and TBRL-LN, but it typically demonstrates 2-5 percentage points lower median 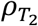 with higher variability and numerous outliers. In S-9 with complex decision rules, G-estimation generates the poorest 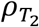 among all methods. TBRL-RF demonstrates consistently inferior performance and generates lower median 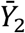, reduced 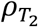, larger *θ*^*^ biases, and the highest variability among all approaches. This performance persists even after calibration that removes up to 96% invalid cutoffs for GO-SMART 40 in S-1. It may reflect TBRL-RF’s dependence on sufficient splitting variables to construct informative and stable decision trees. Random forest constructs trees through bootstrap resampling and random feature selection at each split, which typically improves prediction in high-dimensional settings by reducing overfitting. However, our study only utilizes *Y*_1_ and *d*(*T*_1_, *T*_2_) as splitting variables. Notably, TBRL-RF performs better at avoiding misidentification of nonexistent cutoffs in S-8 while parametric methods like Q-learning tends to identify spurious cutoffs through overfitting to random variation. All analysis methods perfectly identified treatment A as 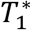.

## 6. Discussion

RA-TF-SMART represents a novel SMART design that integrates RAR based on continuous outcomes into sequential treatment decision-making. It incorporates both within-patient and between-patient adaptation, fully utilizes information available from continuous outcomes to reflect the magnitude and distribution of patient responses. We develop percentile-based and Z-score RA-TF specifications and compare their performance with BR-SMART, TF-SMART and GO-SMART across various trial settings using multiple analytical methods. Our simulation studies reveal that RA-TF-SMART designs consistently achieve superior trial-level outcomes while maintaining statistical validity for post-trial estimation of optimal DTRs. By addressing major limitations of earlier SMART designs, RA-TF-SMART offers a more flexible framework that enhances both patient benefit and statistical inference reliability, particularly in settings where the true treatment effect structure is unknown at trial initiation.

The RA-TF-SMART design offers advantages over BR-SMART and TF-SMART designs in both ethical and practical perspectives. First, RA-TF-SMART randomizes more patients to superior treatment sequences to benefit trial participants, which may enhance trial recruitment and retention.^33,53,54^ Meanwhile, RA-TF-SMARTs eliminate the dependence of TF-SMART on absolute outcome values within pre-specification of outcome ranges. Compared to GO-SMART, our RA-TF-SMART design enhances both the practical implementation of RAR and statistical inference. First, it avoids sensitivity to arbitrary dichotomization cutoffs. The dichotomization in GO-SMART treats outcomes near the cutoff as categorically different regardless of potentially unmeaningful differences caused by measurement error. In practice, identifying an optimal dichotomization cutoff before the trial initiation is difficult because it depends on unknown outcome distributions and treatment effects. Moreover, the performance of GO-SMART critically depends on the adequate cutoff selection to define response indicator. Cutoffs that diverge substantially from the underlying treatment effect structure can force the response rates to be extreme. It may convert modest differences in continuous outcomes into disproportionately large differences in response proportions and further lead to extremely imbalanced sample size across treatment sequences. Hence, GO-SMART may require larger burn-in proportions or sample size to protect against early misallocation. Second, RA-TF-SMART incorporates continuous cutoff updating and EMA smoothing to ensure that randomization probabilities remain appropriate relative to the distribution of outcomes and underlying treatment effect structure. As a result, RA-TFs can generate heterogeneous continuation probabilities in response to true treatment effect cutoffs at different distributional locations. Third, RA-TF-SMART maintains adequate representation across all treatment sequences while providing sufficient adaptation to favor superior treatment sequences, which is essential for robust DTR estimation. GO-SMART forces all responders to continue their first-stage treatment and eliminates exploration of alternative sequences even for patients whose outcomes barely exceed the threshold. Historical controversies regarding the application of RAR in sequential trials focused on potential bias in treatment effect estimation and complications in statistical inference. Our simulation studies reveal that RA-TF-SMARTs offer good statistical power to produce robust post-trial DTR estimates with distributions comparable to BR-SMART and superior to GO-SMARTs.

We develop percentile-based and Z-score RA-TFs since each offers distinct advantages in different trial contexts. Our simulation studies reveal that both methods produce comparable trial outcomes and estimation accuracy in the validation sample. However, Z-score methods slightly optimize the average second-stage trial outcome and proportion of patients whose second-stage treatment assignment matches the optimal DTR, while percentile-based RA-TFs provide slightly better DTR estimations, such as lower proportions of invalid optimal cutoff and more precise optimal cutoff estimates. The percentile-based approach only utilizes the rank information of observed outcomes, making it robust to outliers and non-normal distributions commonly encountered in clinical data. However, if early first-stage trial completers display a narrow range of outcomes, percentile-based cutoffs may not accurately represent the broader population distribution. On the other hand, the Z-score approach prioritizes efficiency by considering the actual magnitude of differences between outcomes under parametric assumption and adjusts second-stage randomization probabilities more aggressively. This performance trade-off has implications for trial design and sample size planning. Implementation of Z-score RA-TF may require larger sample size to establish stable distributional estimates and compensate for higher invalid cutoff rates. Additionally, Z-score methods should be considered if prior data on the outcome of interest suggests a normal distribution. When pilot data or prior studies suggest substantial skewness or numerous outliers, the robust percentile-based methods become more compelling despite offering modestly lower trial-level performance. The choice of RA-TFs should also reflect trial objectives. Z-score approach is preferred when the trial aims to maximize benefit for enrolled participants in a pragmatic effectiveness study, while percentile-based methods outperform if the trial aims to inform future treatment guidelines for earlier-phase treatment development or novel treatment sequences.

RA-TF-SMART may not provide benefits when treatment effects are minimally differentiated across treatment sequences, or when the relationship between stage-one outcome and treatment sequence is non-monotonic or nonlinear. In these settings, RA-TFs may offer limited advantage over balanced randomization or even result in misleading signals. Investigators should carefully consider whether preliminary data or clinical knowledge suggests adequate relationships between stage-one outcome and treatment sequence before implementing RA-TF-SMARTs.

In addition, our simulations assume linear relationships between stage-one outcome and treatment sequence, which favors Q-learning and TBRL-LN since their linear specifications match the true data structure. When outcome relationships are uncertain in practice, TBRL-RF may prove advantageous if we collect sufficient baseline or intermediate covariates. Investigators should also conduct sensitivity analyses across multiple analytical approaches and apply diagnostic tools to assess model specification adequacy as data accumulate. Another important factor to consider is sample size. Our simulations employ 300 patients per trial, which reflects typical enrollment constraints in clinical trial settings due to limited patient populations, restricted resource, or recruitment challenges. Trials enrolling fewer than 200 patients may struggle with simultaneously achieve adequate burn-in representation across treatment sequences and maintain sufficient post-adaptation sample sizes for stable DTR estimation. One limitation of our framework is that it does not incorporate patient baseline or intermediate characteristics. Imbalanced distributions of prognostic factors between treatment sequences may lead to inflated Type-I error and reduced statistical power, especially in RA-TF-SMART designs where RAR intentionally creates unequal allocation probabilities.^32^ Future extensions should investigate covariate-adjusted randomization mechanisms and analytical methods to maintain balance in patient characteristics while preserving the adaptive allocation benefits. Additionally, formal sample size calculation methods for RA-TF-SMARTs should be developed. Also, while this study focuses on RA-TF-SMART designs with continuous outcomes following normal distributions, many clinical outcomes show substantial skewness. In future studies, we aim to extend RA-TFs to accommodate skewed outcomes and apply analytical methods such as generalized linear models and generalized additive model.

In summary, RA-TF-SMARTs represent a promising design to conduct patient-centered sequential treatment randomization that can improve treatment allocation during trials while maintaining the ability to identify the optimal DTR. This design successfully balances the competing goals of maximizing benefit for trial participants and generating generalizable scientific knowledge to guide future trial implementation, making it well-suited for pragmatic trials in real-world clinical settings.

## Data Availability

All data used in the present study were generated through simulation. The code used to generate the data and reproduce the results is available from the corresponding author upon reasonable request.

## Supplementary Materials

**Supplementary Table 1:** Simulation parameters for expected second-stage outcomes across nine scenarios. *β*s and *γ*s denote the treatment-sequence-specific intercept and slope coefficients, respectively.

| Scenario | $\beta_{AA}$ | $\gamma_{AA}$ | $\beta_{AC}$ | $\gamma_{AC}$ | $\beta_{AD}$ | $\gamma_{AD}$ | $\beta_{BB}$ | $\gamma_{BB}$ | $\beta_{BC}$ | $\gamma_{BC}$ | $\beta_{BD}$ | $\gamma_{BD}$ |
| --- | --- | --- | --- | --- | --- | --- | --- | --- | --- | --- | --- | --- |
| S-1 | 20 | 1.2 | 50 | 0.2 | 40 | 0.1 | 10 | 1.2 | 35 | 0.2 | 25 | 0.1 |
| S-2 | 20 | 1.2 | 60 | 0.2 | 45 | 0.1 | 10 | 1.2 | 40 | 0.2 | 30 | 0.1 |
| S-3 | 20 | 1.2 | 30 | 0.95 | 25 | 0.8 | 10 | 1.2 | 18 | 0.95 | 14 | 0.8 |
| S-4 | 20 | 1.2 | 60 | 0.2 | 55 | 0.15 | 10 | 1.2 | 40 | 0.2 | 35 | 0.15 |
| S-5 | 20 | 1.2 | 60 | 0.2 | 20 | 0.1 | 10 | 1.2 | 40 | 0.2 | 10 | 0.1 |
| S-6 | 20 | 1.2 | 60 | 0.2 | 40 | 0.2 | 10 | 1.2 | 40 | 0.2 | 25 | 0.2 |
| S-7 | 20 | 1.2 | 70 | 0.2 | 50 | 0.1 | 10 | 1.2 | 45 | 0.2 | 30 | 0.1 |
| S-8 | 40 | 1.2 | 20 | 0.7 | 30 | 0.8 | 20 | 1.2 | 5 | 0.7 | 10 | 0.8 |
| S-9 | 10 | 1.2 | 32.5 | 0.7 | 47.5 | 0.2 | 5 | 1.2 | 22.5 | 0.7 | 32.5 | 0.2 |

**Supplementary Figure 1.**
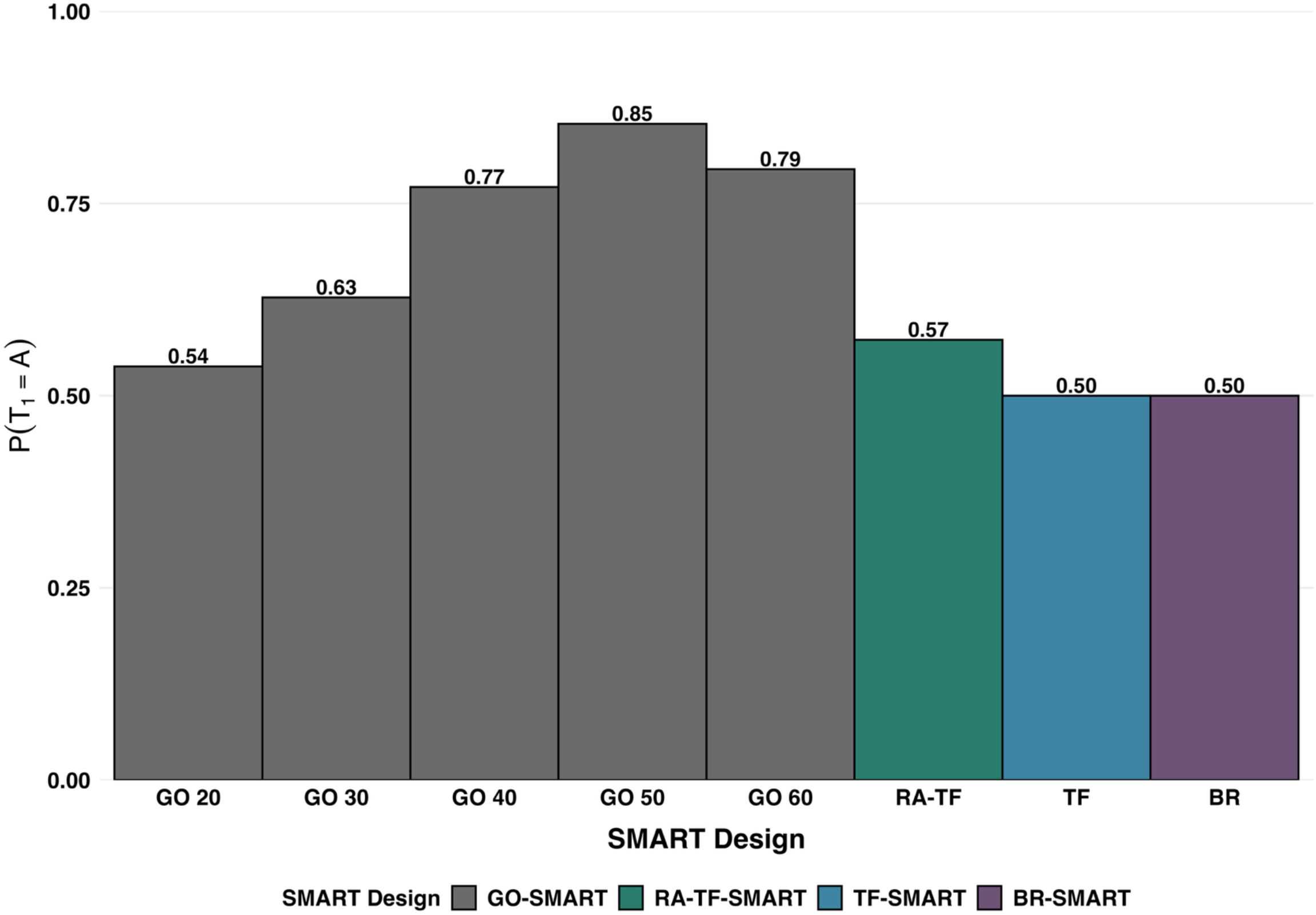
Proportion of patients randomized to the superior first-stage treatment A across sequential multiple assignment randomized trial (SMART) designs. Abbreviations: GO 20 through GO 60, generalized outcome-adaptive SMART with stage-one response cutoffs of 20 to 60; RA-TF, response-adaptive tailoring function SMART; TF, tailoring function SMART; BR, balanced randomization SMART.

**Supplementary Figure 2.**
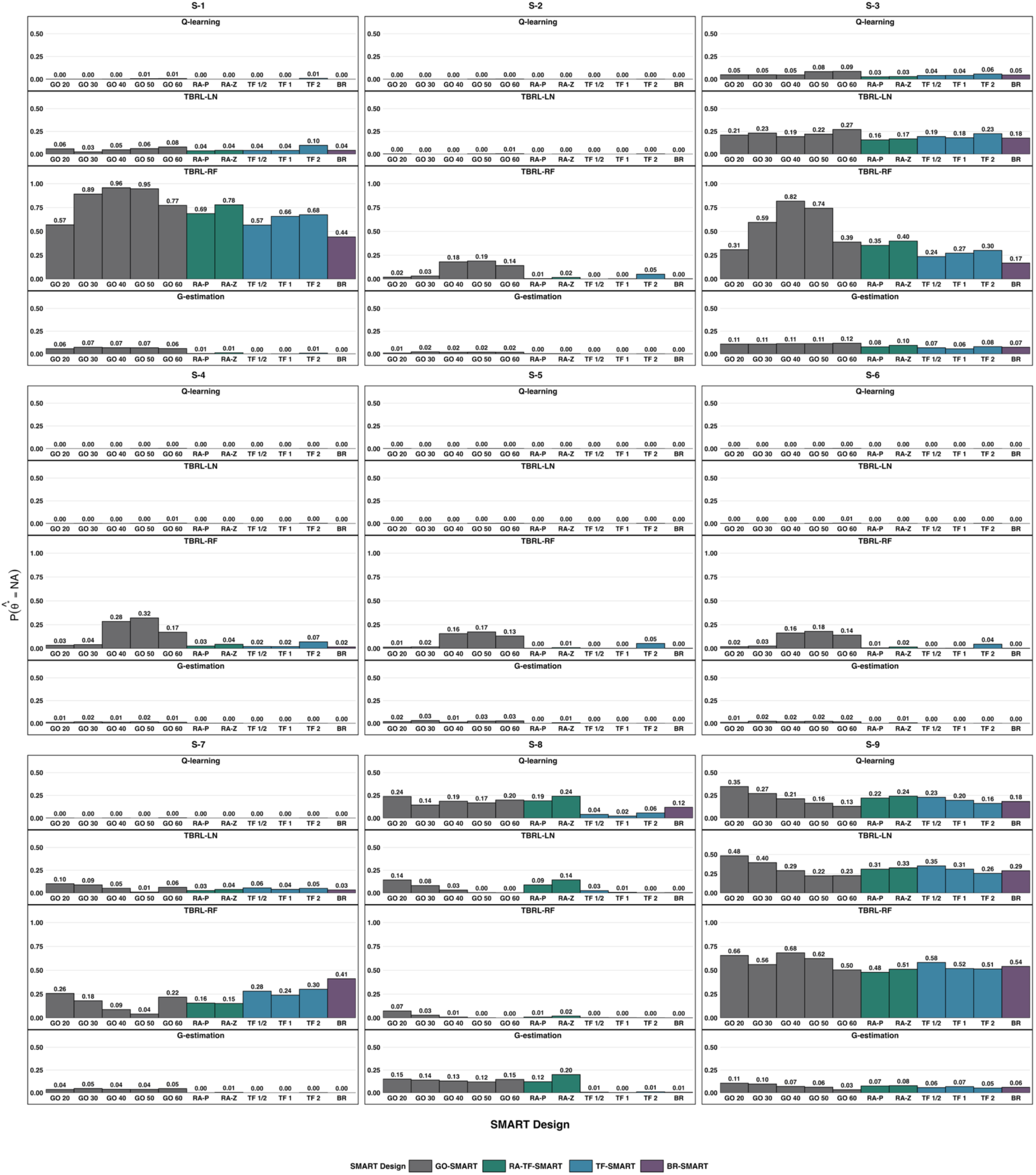
Proportion of simulations with invalid stage-two cutoff estimates across scenarios, sequential multiple assignment randomized trial (SMART) designs, and analytical methods. Abbreviations: TBRL-LN = tree-based reinforcement learning with weighted linear regression; TBRL-RF = tree-based reinforcement learning with random forest; GO 20 through GO 60, generalized outcome-adaptive SMART with stage-one response cutoffs of 20 to 60; RA-P, percentile-based response-adaptive tailoring function SMART; RA-Z, Z-score response-adaptive tailoring function SMART; TF-½, TF-1, and TF-2, tailoring function SMART with adaptation intensity parameters of ½, 1, and 2; BR, balanced randomization SMART.

